# Pre-diagnosis plasma cell-free DNA methylome profiling up to seven years prior to clinical detection reveals early signatures of breast cancer

**DOI:** 10.1101/2023.01.30.23285027

**Authors:** Nicholas Cheng, Kimberly Skead, Althaf Singhawansa, Tom W. Ouellette, Mitchell Elliott, David W. Cescon, Scott V. Bratman, Daniel D. De Carvalho, David Soave, Philip Awadalla

**Author notes:** Corresponding author: Philip Awadalla.

## Abstract

Profiling of cell-free DNA (cfDNA) has been well demonstrated to be a potential non-invasive screening tool for early cancer detection. However, limited studies have investigated the detectability of cfDNA methylation markers that are predictive of cancers in asymptomatic individuals. We performed cfDNA methylation profiling using cell-free DNA methylation immunoprecipitation sequencing (cfMeDIP-Seq) in blood collected from individuals up to seven years before a breast cancer diagnosis in addition to matched cancer-free controls. We identified differentially methylated cfDNA signatures that discriminated cancer-free controls from pre-diagnosis breast cancer cases in a discovery cohort that is used to build a classification model. We show that predictive models built from pre-diagnosis cfDNA hypermethylated regions can accurately predict early breast cancers in an independent test set (AUC=0.930) and are generalizable to late-stage breast cancers cases at the time of diagnosis (AUC=0.912). Characterizing the top hypermethylated cfDNA regions revealed significant enrichment for hypermethylation in external bulk breast cancer tissues compared to peripheral blood leukocytes and breast normal tissues. Our findings demonstrate that cfDNA methylation markers predictive of breast cancers can be detected in blood among asymptomatic individuals up to six years prior to clinical detection.

## Background

High morbidity and mortality rates associated with cancers is largely attributed to late-stage diagnoses. Across most cancers, survival outcomes are significantly improved when tumours are still localised to the tissue of origin at diagnosis [1]. However, effective population screening tools for early cancer detection are currently limited to a few cancer types, notably breast, colorectal, lung and cervical cancer [2,3]. For example, routine mammogram screening is currently recommended to women biennially between the ages of 50-70 in Canada and remains the gold standard for early breast cancer (BRCA) detection. Yet, breast cancer is still expected to be responsible for 25.4% of female cancer cases, and 13.8% of all female cancer related deaths in 2022 [4]. Likewise, limited participation as well as high false positive rates, have raised concerns of overdiagnosis and overtreatment of breast cancers following mammography [5-7].

Profiling cell-free DNA (cfDNA) derived from tumours in blood, also known as circulating tumour DNA (ctDNA), is well demonstrated to be a potential non-invasive biomarker that can provide a glimpse into the genetic and epigenetic landscape of a tumour’s genome [8-12]. Sensitive liquid biopsy assays examining tumour specific cfDNA methylation profiles are able to detect both early- and late-stage cancers and inform on the tissue of origin of underlying tumours. In addition, some studies have even combined cfDNA biomarkers with alternative markers such as multi-protein panels or radiographic imaging to further improve diagnostic accuracy [11, 13]. Several studies to date have shown the diagnostic potential of cfDNA methylation profiles for early breast cancer detection by building targeted panels from bulk cancer tissue to profile and classify individuals with established early-stage cancers [13-16]. However, as most cancers are often only detected once patients are screened or become symptomatic, these studies have primarily been performed using biologic samples collected from patients following clinical detection and diagnosis of a malignant primary tumour. Profiling cfDNA in the pre-diagnostic context could allow us to better understand the detectability of cancer biomarkers across cancer subtypes at the earliest stages, however this requires application of new technologies to biologics collected from healthy individuals prior to a cancer diagnosis.

Here, we profiled cfDNA methylation patterns in plasma samples collected from cohort participants prior to a breast cancer diagnosis and matched cancer-free controls to identify and assess cfDNA markers predictive of early breast cancers and breast cancer risk. We leverage the Ontario Health Study (OHS), an Ontario-based longitudinal prospective cohort that collected health and lifestyle information through self-reported questionnaires, and biologics including blood plasma, from over 41,000 participants between 2009 and 2017 [17]. A particular advantage of the OHS is that almost all participants provided consent to administrative health linkages at initial recruitment into the study. We were able link health insurance numbers of recruited individuals to administrative health registries to identify participants that developed breast cancer up to 7 years after study recruitment and biologic donation. Using 1.6 mL of blood plasma from incident breast cancer cases, in addition to matched controls, we analyzed and compared cfDNA methylomes in pre-diagnosis blood plasma samples versus cancer-free samples. In this study, all sequencing runs and analytics in the discovery cohort were performed with cases and controls concurrently to minimize inflation of accuracy, sensitivity, and specificity. By retrospectively interrogating blood samples collected prior to diagnosis, we assessed the earliest detectability and predictive performance of cfDNA methylation markers for classifying participants harboring undetected breast cancers and in stage IV breast cancers from an independent cohort.

## Methods

### Cohort participants and demographics

Peripheral blood was drawn from OHS participants upon recruitment to the study, and 1.6 mL plasma was separated and collected within 48 hours into EDTA tubes, and immediately cryopreserved at the OHS Biobank. Participants in OHS that had developed breast cancer following recruitment to the study were identified by linking individuals to the Ontario Cancer Registry through Cancer Care Ontario (CCO) using health insurance number, age, sex and name. At the time of linkages, cancer registry data had been made available through the Ontario Cancer Registry up until December 2017. All samples and participant data were deidentified and assigned unique research IDs to prevent identification of study subjects prior to analysis. Original OHS and CCO IDs are not known to anyone outside the research group. Breast cancers were confirmed by histological analyses of tissue biopsies at the time of diagnosis, and immuno-histochemical tests for hormone receptor status were reported in the pathology records of breast cancer cases. In total, 110 incident breast cancer cases among participants diagnosed with breast cancer after providing a blood sample at the time of enrollment were identified, in addition to 108 control participants with no history of cancer at the time of study enrollment and throughout the study follow up time (Fig. 1, Fig. S1, Supplementary Table 1 & Supplementary Table 2). Cancer-free controls were matched to cases by age, sex, date of biologic collection, ethnicity, smoking status, and alcohol consumption frequency were also selected. Additional plasma samples from 35 patients with established breast cancer were obtained from participants in the Ontario-wide Cancer TArgeted Nucleic Acid Evaluation (OCTANE) study [18].

**Fig. 1:**
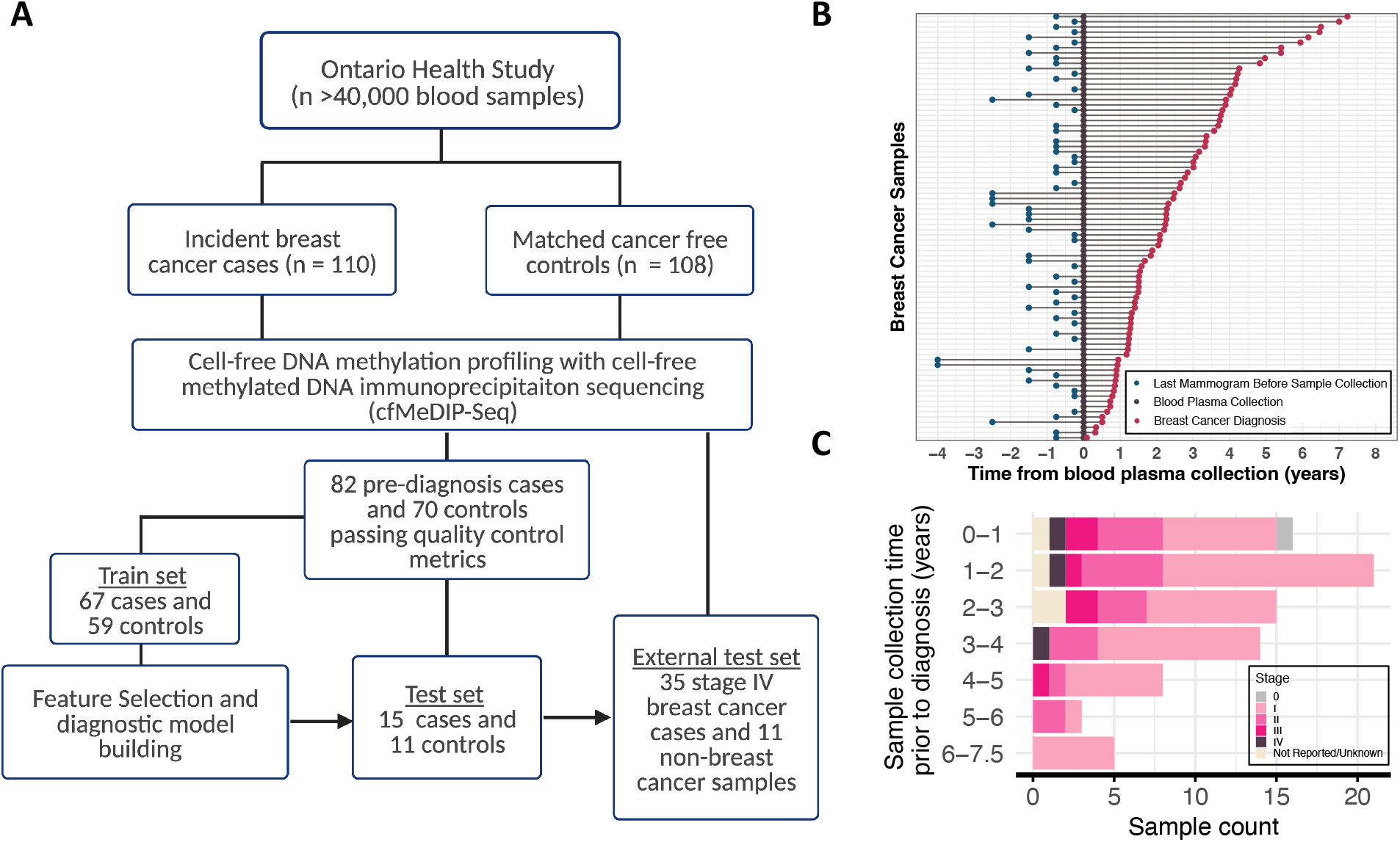
Overview of study design and incident breast cancer cases in the Ontario Healthy Study. **(A)** Outline of participant recruitment and blood plasma sample selection process in the Ontario Health Study (OHS). Cell-free DNA methylome of blood plasma from 218 OHS participants profiled with cell-free methylation DNA immunoprecipitation sequencing (cfMeDIP-Seq). 152 samples passed all quality control metrics. Train set of 67 pre-diagnosis breast cancer cases and 59 cancer-free controls were used to identify differentially methylated regions for building breast cancer diagnostic classifiers. Held-out batch of 15 pre-diagnosis breast cancer cases and 11 cancer-free controls used to evaluate diagnostic classifier predictive performance. External test set of 35 breast cancer samples collected at diagnosis time and 11 non-breast cancer samples were used to further validate diagnostic classifier performance. **(B)** Timeline of blood plasma collection, breast cancer diagnosis and last mammogram prior to biologic collection across incident breast cancer cases in OHS **(C)** Pre-diagnosis OHS cases across time between sample collection and breast cancer diagnosis. Colors indicate stage at diagnosis across cases.

### Cell-free DNA methylation profiling

Using 1.6 mL of plasma from pre-diagnosis breast cancer and selected cancer-free control participants, cfDNA methylation patterns were profiled using a cell-free methylated DNA immunoprecipitation sequencing protocol (cfMeDIP-Seq) to sequence methylated cfDNA fragments [19-20]. The cfDNA from OHS and OCTANE samples was extracted from plasma using the QIAamp Circulating Nucleic Acid Kit (Qiagen). 5-10ng of cfDNA was used as input to generate methylated cfDNA libraries (IP libraries) along with an input control library (IC libraries). Quality of incoming cfDNA was assessed using the Fragment Analyzer (Agilent) following the manufacturers guidelines. 0.1ng of *Arabadopsis thaliana* DNA was added to samples prior to library preparation. Combined samples were prepared using the KAPA Hyper Prep library protocol (Roche), with standard End Repair & A-tailing and ligation of xGen Duplex Seq Adapter (IDT), followed by incubation at 4°C overnight. Unmethylated lambda (λ) DNA was added to partially completed IP libraries and enriched for methylated DNA using the MagMeDip Kit (Diagenode) and purified with the IPure Kit v2 (Diagenode). Sample indices were added to IP and IC libraries via PCR. Completed libraries were quantified by Qubit (Life Technologies) and Fragment Analyzer (Agilent). Both IP and IC libraries underwent shallow sequencing (∼20,000 reads) on the MiSeq platform as a quality control step. IP libraries were sequenced to approximately 60M read pairs in 2×50bp mode on Novaseq platform (Illumina). To mitigate confoundment of biological signals from technical artifacts associated with batch effects, cancer cases were batched together with control samples during and across library preparation and sequencing runs.

### Raw sequencing read processing

Following sequencing, the FASTQ raw reads were adapter trimmed, with unique molecular identifiers (UMIs) appended to fastq headers using UMI Tools (version 0.3.3) [21]. The reads were then aligned to hg38 using Bowtie2 (version 2.3.5.1) [22] in paired end mode at default settings. Aligned SAM files were converted to BAM file format, indexed, and sorted using SAM tools (version 1.9) [23]. Aligned reads were subsequently deduplicated according to alignment positions and UMIs using UMI Tools.

### Quality control and sample inclusion

One control sample was excluded from our study owing to mortality from non-cancer related causes during study follow up. Five controls were excluded due to diagnoses of cancer pre-disposing conditions that were identified from self-reported questionnaires during follow up. Three control samples were excluded owing to diagnosis of another cancer following sample collection and processing. Following library preparation, four samples were removed as no reads were generated during the MiSeq quality control step. We retained and analysed all samples with more than 10 million deduplicated reads. 39 samples were removed owing to Novaseq sequencing instrument failure that resulted in poor sequencing yields. To assess enrichment efficiency, the number of methylated and unmethylated Arabidopsis spike-ins aligned to F19K16 and F24B22 respectively were counted, and the proportion of methylated spike-ins generated out of the total spike-ins were calculated. Seven samples with less than 95% of spike-in reads that were methylated were excluded. An additional seven were samples owing to poor CpG enrichment assessed through GoGe (< 1.75) and relH enrichment scores (< 2.7) calculated using MEDIPS were also removed (See Supplementary Table 2 for quality control metrics and sample information among remaining samples). Following quality control filtering, 82 pre-diagnosis breast cancer cases and 70 cancer-free controls were retained for subsequent analyses. Additional previously published cfMeDIP-Seq profiles from six head and neck cancers cases and five non-cancer controls were used as non-breast cancer controls for validation [24].

### Computing cfMeDIP-Seq methylation signals

To infer cfDNA methylation levels among pre-diagnosis breast cancers and control samples, coverage profiles were generated for each sample across 300 bp non-overlapping binned tiled windows from BAM files using MEDIPS (R package version 1.12.0) [25]. Cell-free DNA methylation coverage profiles were library size normalized across all OHS, OCTANE and external non-breast cancer samples using the DESeq2 R package version 1.30.1 [26]. Regions with no coverage within a particular sample are assigned a count of 0. To reduce background noise, publicly accessible data was used to remove potentially uninformative regions. Regions frequently methylated in haematopoietic cells were inferred using whole genome bisulfite sequencing data of peripheral blood leukocytes (n = 78) from the International Human Epigenetics Consortium (IHEC) [27]. We averaged the level of methylation across all CpG sites within the same 300 bp non-overlapping tiled window for each sample to infer the level of methylation within a specified region. Regions with a methylation level greater than 0.25 averaged across PBL samples for each cell type were excluded. Remaining 300-bp bins with at least six or more CpG sites located at CpG islands, shores and shelves, or in FANTOM5 annotated promoters and enhancers, or UCSC RepeatMasker repetitive elements were tested for differential methylation (Fig. S2) [28].

### Statistical analysis

All statistical analyses were implemented in R, version 4.0.4.

### Pre-diagnosis breast cancer differential methylation calling, classifier building and performance assessment

OHS samples were divided into a discovery set (n = 67 cases and n = 59 controls) and validation set (n = 15 cases and n = 11 controls). Discovery set samples were used to identify differentially methylated regions (DMRs) and to build predictive models for classifying early breast cancers. While remaining validation set samples from a held-out batch, processed independently from the discovery cohort samples, were used as a test set to validate the discovery cohort signatures and predictive model.

To evaluate the best approach for differential methylation calling and the optimal number of features to train machine learning models for predicting breast cancer risk among pre-diagnosis samples in discovery set samples, a repeated 10-fold cross validation (CV) was performed 100 times (Fig. S3) [29]. First, pre-cancer cases and control samples were divided into 10 approximately equal sized sets using stratified sampling, balancing the proportion of pre-diagnosis cases by years prior to diagnosis following blood collection in each fold set. Iteratively, for each fold in the CV procedure, one set was selected as the test set and the remaining nine sets were designated as the train set (comprising 10% and 90% of participants respectively). Within the train set folds, differential methylation calling was performed using a Wald test of the coefficient from a negative binomial regression of cfMeDIP-Seq methylation level on train set case and control status using DESeq2, adjusting for batch using surrogate variables and age as covariates. Differential methylation variance was tested by performing Bartlett’s test using matrixTests (R package version 0.1.9.1) [30] between pre-diagnosis cases and controls. Features were ranked according to p-values from respective tests selecting for regions with a minimum log fold-change in methylation > 0.5 between cases and controls.

Iteratively, within each subsampling iteration, the top ranking 50 to 400 hypermethylated regions identified from train set folds were used to construct a random forest model with Caret (R package 6.0) [31, 32] from library-size normalized discovery set sample methylation counts. The random forest models were tuned using a nested 10-fold cross validation with 10-repeats iterating across values from 5 to 50 for the number of features sampled to grow individual decision trees, and 250 to 1000 trees in the model to maximize the overall nested CV classification accuracy. The model performance was then assessed by applying the predictive model to the held-out test fold to obtain classification scores that reflected the proportion of decision trees predicting the sample as breast cancer. The 10-fold CV procedure was repeated 100 times with different fold-splits to get stable average estimates of cross-validated predictive performance. Differential methylation calling, classifier building and assessment of predictive performance was effectively repeated 1000 times with different sets of cases and controls in the train folds across each iteration. The test-fold classification scores across each of the 100 CV repeats were averaged for each sample. A bootstrapped area under the receiver operating characteristic curve (AUC) was calculated by subsampling 67 cases and 59 controls with replacement, repeated for 3000 bootstraps. The final bootstrap AUC was calculated by taking the median AUC, while 95% confidence intervals were calculated by taking 2.5% and 97.5% percentiles.

To incorporate the follow-up time of controls, the time to diagnosis among the pre-diagnosis cancer cases, as well as the proportion of true negative cases in the population, the concordance index (C-index: the probability that for any pair of individuals, the individual with the higher estimated risk score will have the earlier diagnosis time) [33] and time-dependent AUCs (AUC(t)) were computed [34]. Owing to the outcome and age dependent sampling used in the study, the artificial case-to-non-case ratio in our sample was not representative of the Canadian adult population. Sampling weights were calculated to adjust for this sampling bias in the time-dependent model assessment analysis using age specific cumulative breast cancer incidence rates from the Canadian Cancer Registry, in addition to all-cause mortality rates in Ontario reported by Statistics Canada. Kaplan-Meier estimates weighted according to age-specific breast cancer incidence for the corresponding follow-up times in the Canadian population were also computed to assess whether averaged test-fold classification scores were predictive of time to cancer diagnosis. Samples were stratified into a high risk and low risk group according to whether they were assigned a classification score above or below 0.638 respectively. The cut off was determined by optimizing sensitivity while limiting false positive rates at 5%. Cumulative probabilities of developing breast cancer across timepoints were estimated by fitting a Cox Proportional Hazard (CoxPH) model using the *cph* function (rms R package version 6.3) on the discovery set with classification scores as predictors.

### Validation of discovery cohort signatures

To validate the predictive performance of pre-diagnosis cfDNA hypermethylated regions, the mean p-value and log fold-change (logFC) was calculated across the 1000 repeated differential methylation calls within the discovery cohort CV procedure. The top ranking features were used to build a random forest model from all discovery cohort samples and tuned using a nested CV in the same way described above to optimize the tree number and number of subsampled regions for each tree. To infer whether pre-diagnosis cfDNA DMRs were agreeable with established cancers, predictive models were further assessed on late-stage breast cancer cases (n = 35) from OCTANE [18] and external head and neck cancer cases [24] (n = 6), and cancer free controls (n = 5). Predicted risk scores in the validation sets were used to calculate AUROCs to assess classification performance and weighted Kaplan-Meier estimates to assess the performance for predicting the absolute risk for developing breast cancer. The observed cumulative incidence rates were compared to those estimated by the discovery set CoxPH model applied to the validation samples.

### Overlap between pre-diagnosis cfDNA hypermethylated regions and bulk breast cancer tissue methylome

To assess whether the selected predictor regions used to build the classification model were potentially derived from breast cancer tissue, the number of overlapping hypermethylated regions was calculated between pre-diagnosis breast cancer cfDNA and bulk breast cancer tissue relative to adjacent breast normal (ABRNM), healthy breast normal (HBRNM) tissue, and peripheral blood leukocytes (PBL) using publicly accessible 450k DNA methylation array data. Solid breast cancer and normal tissue raw IDAT files were downloaded from the TCGA data portal, and PBL from the GeoExpression Omnibus (GSE87571 and GSE42861), in addition to healthy and adjacent normal tissues (GSE88883, GSE101961 & GSE66313). IDAT files were processed to generate beta methylation values from IDAT files using Minfi (1.36.0 R package) [35] and normalised using the *preprocessFunnorm* function. To test for differentially methylated CpG sites between paired healthy and tumour biopsies, an F-test was performed using the DMPFinder function from Minfi across 485,512 CpG sites. Significantly differentially methylated regions were defined as CpG sites with an absolute difference in methylation of greater than 0.1 and Bonferroni adjusted p-value of less than 0.0001. Differential methylation calling between all breast cancer and breast normal tissue, as well as between breast cancer tissue and PBL was also performed using the *DMPFinder* function from Minfi between all samples from each respective group to identify additional breast cancer specific markers. A permutation analysis was performed to infer whether overlaps were significant, by comparing the observed overlap with the overlap between significantly hypermethylated bulk tissue DMRs and randomly selected background cfDNA regions (repeated 3000) times to obtain a background distribution for z-score normalization and to calculate corresponding p-values. Additional BRCA, ABRNM, and HBRNM tissue data profiled from the same study (GSE69914) with processed Beta methylation values was also analyzed as described above. Predictor regions were annotated for genomic contexts using Annotatr (R package version 1.24.0).

## Results

### Discovery cohort pre-diagnosis breast cancer classification predictive performance; internal cross-validation

Most early cancer detection studies to date typically use a pre-designed enrichment panel to target cancer tissue specific differentially methylated genomic loci prior to methylation profiling [13-15]. In this study, we instead utilized cfMeDIP-Seq to interrogate genome-wide methylation profiles, enabling the detection of both cfDNA specific methylation markers derived from tumours and differentially methylated regions from non-tumour material potentially predictive of breast cancer risk. To reduce background noise, we applied biological filters to remove potentially uninformative genomic regions prior to differential methylation calling (Fig. S2). Loss of epigenetic stability and increased stochasticity across the genome have been observed in pre-malignant and early cancer tissue [36, 37]. Likewise, we suspect that in pre-diagnosis cfDNA samples not all regions hypermethylated in cancer tissues will be observed, nor will we observe consistent genomic regions to be impacted by methylation changes across pre-symptomatic individuals. Therefore, we compared two approaches for differential methylation calling to rank and select regions in the discovery cohort for training predictive models; the Wald test of the negative binomial coefficient to test for changes in mean methylation level between groups, and the Bartlett’s test identifying differential variance in methylation to improve detection of sparse predictor regions.

To iteratively identify and assess, using multivariate predictive models, whether cfDNA hypermethylated regions can accurately predict an individual’s risk of developing breast cancer, we performed 100 repeated 10-fold CV procedures in the discovery cohort (Fig. S3). As the predictive performances can be highly variable depending on which observations are included in the train set and test set, a repeated CV approach can obtain stable average estimates as well as the uncertainty of the cross-validated predictive performance in held-out test folds among discovery cohort samples. In practice, a diagnostic screening tools aims to minimize false positive rates while maximizing sensitivity to prevent overdiagnoses, thus we assessed the optimal number of predictors and best feature selection approach that achieved the highest sensitivity at 95% specificity. Across all CV repeats, random forest classifiers trained with the top 150 hypermethylated regions from the Wald’s test achieved the highest average sensitivity at 26.9% (95% CI 0.9%-49.3%) while retaining 95% specificity for predicting breast cancer development on test-fold samples (Fig. 2, Fig. S4). The averaged classification performance from bootstrapped classification scores in discovery cohort samples achieved a mean binary classification AUC across all breast cancer types, ages and varying pre-diagnosis time intervals of 0.724 (95% CI 0.636-0.810) (Fig. 2A). Further, the diagnostic classifiers trained with top 150 ranking hypermethylated regions achieved an average C-index of 0.704 (95% CI 0.647-0.758) across the repeated 10-fold CV iterations up to seven years prior to diagnosis in test set folds (Fig. 2B). The diagnostic classifiers performed consistently well among cases diagnosed at stage I, achieving a mean AUC of 0.725 (95% CI 0.626-0.824) and mean C-index of 0.704 (95% CI 0.655-0.757) (Fig. 2C). Notable differences in predictive performance across different breast cancer subtypes were also observed when stratifying predictive performance by hormone receptor (HR) positive (n = 43 cases; 64.2%) and HR negative (n = 7 cases; 10.4%) breast cancers. Discovery cohort classifiers performed better identifying early HR positive breast cancer cases using pre-diagnostic blood cfDNA methylation signatures, detecting on average 32.6% of cases (95% CI 8.7%-50%) at 95% specificity compared to an average sensitivity of 14.3% (95% CI 0%-43.2%) for HR negative cancers (Fig. 2C).

**Fig. 2:**
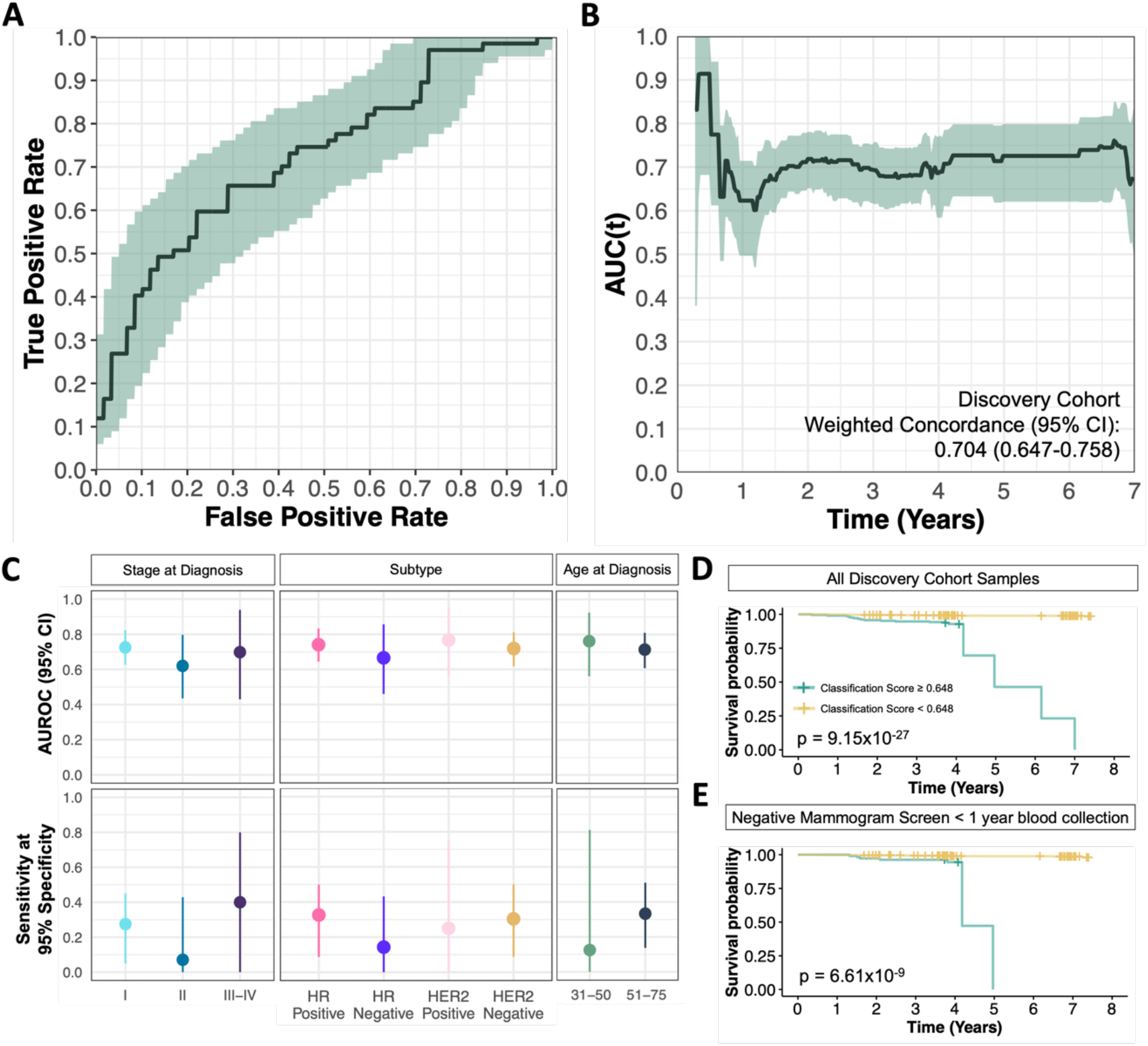
Classification performance of discovery set OHS pre-diagnosis cases and controls using top 150 hypermethylated regions. Test-fold classification scores averaged across 100 repeats for each sample. **(A)** Bootstrapped receiver operating characteristic (ROC) curves of discovery cohort sample classification scores. Mean performance across 1000 bootstraps are shown in black, with 95% confidence intervals indicated by shaded green regions. **(B)** Time-dependent area under ROC curves (AUC(t)) weighted for the true cumulative age-specific breast cancer incidence rates in Canada. Mean AUC(t) shown in black lines, with shaded regions indicating 95% confidence intervals **(C)** Bootstrap AUROC and sensitivity at 95% specificity for classifying discovery cohort stratified by subtype at diagnosis, stage at diagnosis, and age at diagnosis. Dots show mean performance, while lines indicate 95% confidence intervals **(D&E)** Kaplan-Meier curves indicating cancer-free survival time following blood collection across **(D)** all samples and **(E)** in samples with a negative screening mammogram within one year of blood collection. Samples are stratified by mean classification score above or below 0.648.

Typically, all women between the ages of 50-70 are recommended to receive mammograms biennially in Canada, however mammographic screening guidelines for women aged 40-49 remains controversial as The Canadian Task Force on Preventive Health Care recommended against routine screening of women under 50 in their 2018 guidelines [7]. To address the differences in routine care within the cohort, we specifically evaluated whether individuals diagnosed at ages 35 to 50 in the discovery set, preceding the age of mammographic screening eligibility in Ontario, could also benefit from cfDNA methylation tests for early breast cancer detection. When stratifying binary classification performance according to age of diagnosis, individuals diagnosed between the ages 35 to 50 (n = 16 cases) were classified with an AUC of 0.775 (95% CI 0.624-0.895), detecting 25% (95% CI 6.2%-62.5%) at 95% specificity, and a C-index of 0.743 (95% CI 0.633-0.850) (Fig. 2C). Furthermore, within the discovery cohort, 35 cases reported to have a negative breast mammography screen result within six months to one year before providing a blood sample to the OHS (Fig. 1B). Stratifying the classification performance for cases with negative mammogram results within one year of providing blood samples (Fig. 2C) reveals classifiers achieve an AUC of 0.691 (95% CI 0.581-0.803), and a 20% sensitivity (95% CI 5.7%-41.0%) at 95% specificity, highlighting that cfDNA methylation markers can be predictive of breast cancers prior to mammogram detection.

The average classification scores assigned by predictive models in each cross-validated fold of the 100 repeats were also highly predictive of cancer-free survival. Due to the inflated ratio of cases to controls in our study, unweighted KM curves will report inaccurate cancer-free survival probabilities, particularly in low-risk groups as the proportion of cases to controls is significantly lower in OHS and across the Canadian population (Fig. S5B-E). As such, we weighted case and control samples by the age specific cumulative breast cancer incidence rates, adjusted for the all-cause mortality rates in Ontario. Using a classification score cut off of 0.648 to stratify samples using a weighted Kaplan-Meier (KM) estimate, which retains 95% specificity among discovery cohort samples (Fig. S5A), we demonstrate significant association between high classification scores and cancer-free survival rates (log-rank test p = 9.15×10^−27^) particularly in HR positive breast cancers cases (log-rank test p = 9.70×10^−27^) and cases diagnosed between ages 35 to 50 (log-rank test p = 3.4×10^−31^) (Fig. 2D-E & Fig. S5G-H). Similarly, when estimating the cumulative incidence rate of developing breast cancer within varying time points between high risk (classification score ≥ 0.648) and low risk (classification score < 0.648) groups using a CoxPH model, we found that individuals with a high risk score had on average an 11.1% (5.7%-16.1%) chance of developing breast cancer within five years (Supplementary Table 3). Indeed, our observed cumulative incidence was consistent with our estimated incidence at up to 4 years, after which the observed incident was significantly higher owing to limited control samples with long censorship times being predicted in the high risk group. Collectively, our findings reveal that hypermethylated cfDNA signatures identified from pre-diagnosis samples can be predictive of breast cancer development in our discovery cohort.

### External test set validation of discovery cohort differentially methylated regions and breast cancer diagnostic classifier

To validate whether pre-diagnosis cfDNA hypermethylated regions could predict the risk of an individual developing breast cancer, we built a random forest classifier trained using all discovery cohort samples and assessed on the held-out validation set of pre-diagnosis cases and controls that were processed independently from samples in the discovery set. As the top 150 hypermethylated regions from the Wald’s test achieved the highest average sensitivity in the discovery set across internal CV repeats, we selected the same number of top ranking features to build a classifier to predict breast cancer development in the OHS validation set samples.

Predictor regions used to train the model were selected by ranking hypermethylated regions according to the mean p-value from the Wald’s test across the 1000 repeated subsampling differential methylation calls for each region within the discovery set. The diagnostic classifier trained using the top 150 hypermethylated regions accurately discriminated validation set pre-diagnosis breast cancer cases from controls, achieving an AUC of 0.930 (0.815-1.000) among pre-diagnosis test set samples. Using the classification score cut off of 0.648, determined previously from discovery set samples, we classified 53.3% of samples at a 0% false positive rate (Fig. 3A-C). Test set sample classification scores assigned by the diagnostic classifiers were also significantly higher (p = 4.1×10^−5^) in pre-diagnosis cases relative to controls (Fig. 3B). Interestingly, all test set OHS samples with a classification score of over 0.648, selected previously from discovery cohort samples, developed breast cancer within 6 years of blood sampling, demonstrating cfDNA methylome signatures can be predictive of breast cancer risk (Fig. 3D). Using the CoxPH model fitted from the discovery set, samples in the validation set high risk group had an average estimated 9.2% (4.7%-13.4%) chance of being diagnosed with breast cancer within 5 years. Owing to limited sample sizes, the average incident rate among high risk group samples was underestimated compared to the observed incident rate, which were considerably higher as no validation set control samples were assigned in the high risk group. Consequently, further examination in a larger cohort is needed to assess the calibration of estimated risk incidence.

**Fig. 3:**
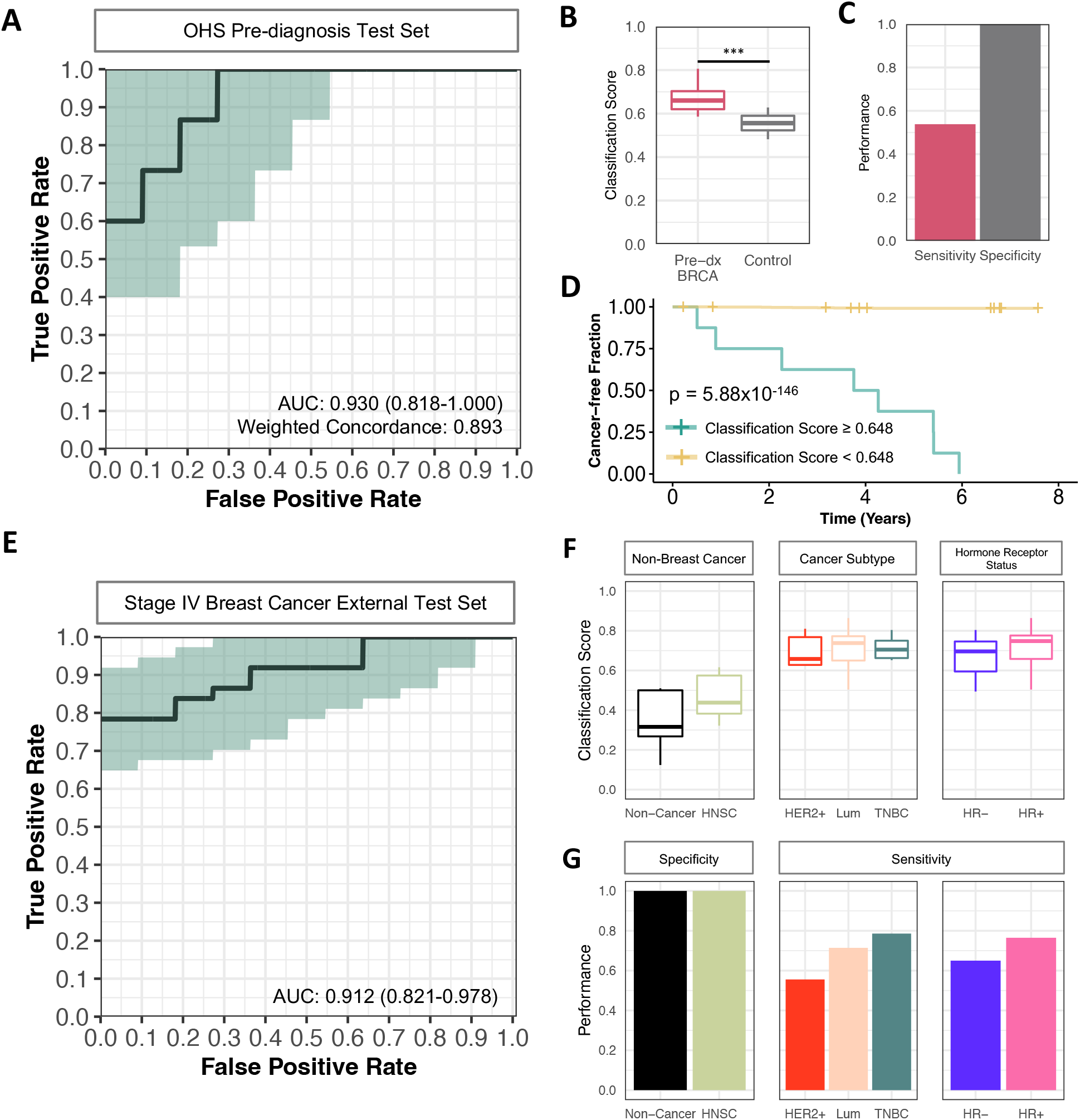
Classification performance of independent validation set pre-diagnosis and late-stage breast cancer cases and controls. **(A)** ROC for classifying test set pre-diagnosis breast cancer cases (n = 15) and controls (n = 11) from a separate batch of OHS samples. Predictive performance of classifier trained using top 150 ranked hypermethylated regions identified from discovery cohort samples **(B)** Predicted classification score among OHS test set pre-diagnosis cases and controls (p = 4.10×10^−5^) and **(C)** corresponding sensitivity and specificity using a classification cut off of 0.648 previously determined from discovery cohort samples. **(D)** Kaplan-Meier survival curves of pre-diagnosis test set samples grouped by classification score cut off above (green) or below (yellow) 0.648 (log-rank test p = 5.88×10^−146^). Cut off determined from discovery cohort samples that yielded 95% specificity. **(E)** ROC for classifying all external test set of metastatic breast cancer cases collected at the time of diagnosis (n = 35) cases and non-breast cancer controls (n = 11). External samples were classified by the diagnostic model built from discovery cohort samples. Samples from breast cancer cases were collected at the time of breast cancer diagnosis before treatment **(F&G)** External test set classification scores across non-cancer controls (n = 5), head and neck squamous cell carcinomas (n = 6) cases, and late-stage breast cancer (n = 35) samples. **(F)** Classification scores of external test set samples from diagnostic classifier stratified by non-breast cancer samples, PAM50 and hormone receptor subtype **(G)** Specificity and sensitivity across breast cancer subgroups for classifying external test set samples using cut-off score of 0.648 to discriminate breast cancer cases from controls.

We further evaluated the performance of the diagnostic classifier on an independent test set comprised of stage IV breast cancer cases (n=35) profiled at the time of enrollment into OCTANE, head and neck squamous cell carcinoma (HNSC) cases (n=6), and a set of non-cancer controls (n=5) from samples profiled by *Burgener et al* [18, 24]. The classifier achieved an AUC of 0.912 when predicting on individuals with established late-stage breast cancer against non-breast cancer samples regardless of subtyping (Fig. 3E). Interestingly, the classifiers performed better in detecting HR positive breast cancers (AUC=0.963) compared to HR negative (AUC=0.868) among the external test samples, consistent with the discovery cohort classification performance. Using a classification score cut-off of 0.648, the classifiers detected HR positive breast cancers at 76.4% sensitivity and HR negative breast cancers at 65.0% sensitivity, while retaining a specificity of 100% for non-breast cancer samples including head and neck cancer cases (Fig. 3F-G), indicating that the pre-diagnostic breast cancer cfDNA methylation signatures are largely agreeable with late stage malignancies.

### Differentially methylated regions in pre-diagnosis breast cancer cfDNA reflects breast cancer epigenome

We next assessed whether the top 150 ranked cfDNA hypermethylated predictor regions identified from discovery cohort samples were concordant with breast cancer tissue hypermethylated regions. However, as participant tumour biopsies at the time of diagnosis were not available, we instead leveraged publicly available bulk breast cancer, adjacent breast normal, healthy breast normal and PBL DNA 450k methylation array data. Presumably, if the cfDNA hypermethylated markers were derived from breast cancer tissue, the same regions would be concordantly hypermethylated when comparing bulk breast cancer tissue methylomes against PBLs, as cfDNA from normal breast tissue is typically shed at extremely low frequency in healthy individuals. Across the top 150 pre-diagnosis breast cancer cfDNA hypermethylated regions, 75 regions contained at least one CpG site profiled by the DNA 450k methylation array spanning 156 CpG Sites. The difference in methylation between TCGA bulk breast cancer tissues (n = 846) and PBLs (n = 628) across the 156 CpG sites overlapping the 75 cfDNA hypermethylated regions were computed against the cfDNA log-fold change in methylation (Fig. 4A), revealing that hypermethylated regions in pre-diagnosis breast cancer cfDNA were concordantly hypermethylated in bulk breast cancer tissue relative to PBLs. Similarly cfDNA hypermethylated regions can also discriminate bulk breast cancer tissue from PBLs and bulk breast normal tissues (Fig. S6).

**Fig. 4:**
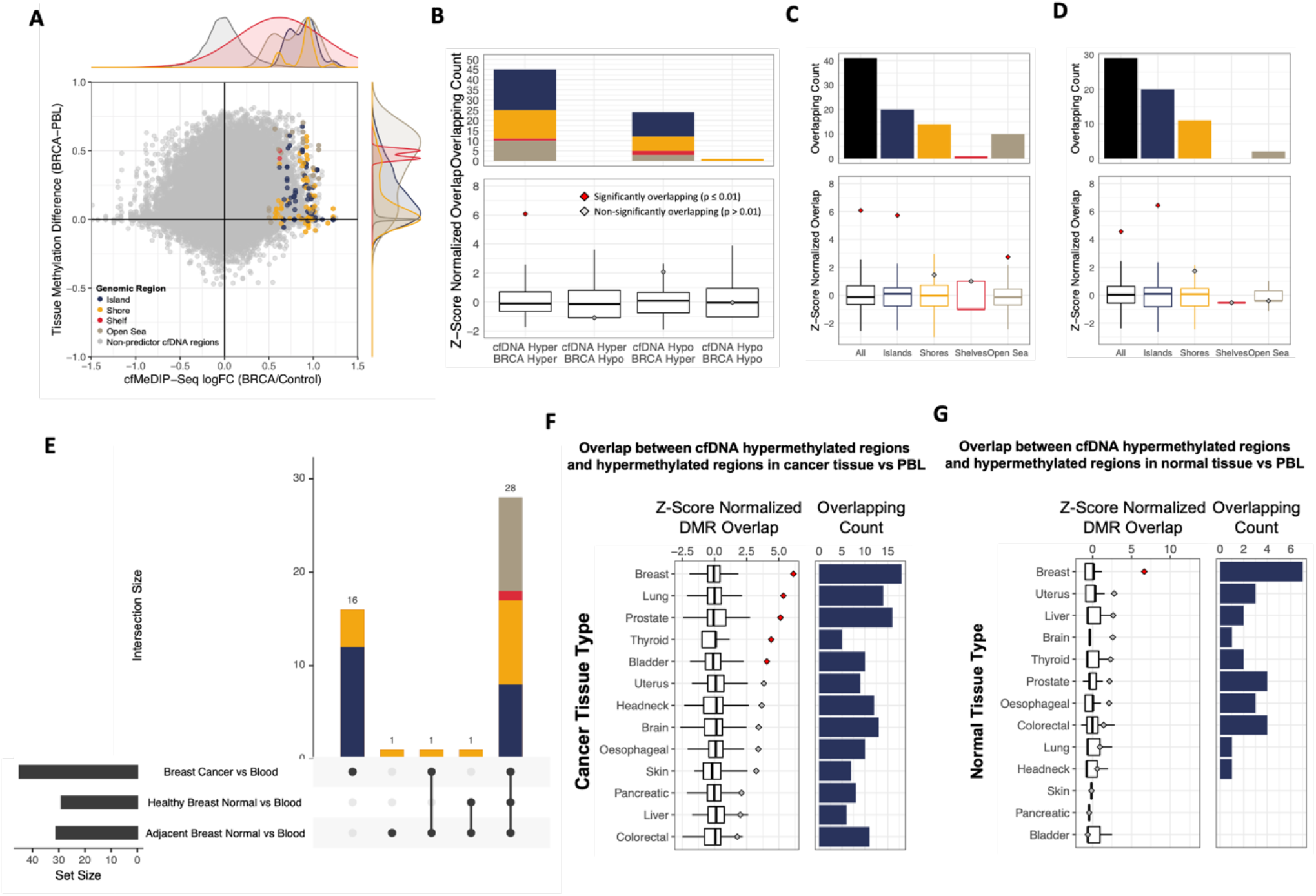
Overlap between top 150 pre-diagnosis breast cancer cfDNA hypermethylated regions and bulk tissue hypermethylated regions. **(A)** Association between the log fold-change in cfMeDIP-Seq cfDNA methylation of discovery set pre-diagnosis cases versus controls and difference in 450k array beta methylation level between bulk breast cancer tissue versus PBLs. Each point represents a cfDNA region overlapping a 450k methylation array CpG Site. Gray dots represent background cfDNA regions, while non-grey colored points represent CpG sites in 75 out of the top 150 differentially hypermethylated regions in pre-diagnosis cfDNA colored by CpG islands (blue), shores (yellow), shelves (red) or open sea (brown) regions. **(B)** Number of overlapping regions between the top 150 pre-diagnosis breast cancer cfDNA DMRs, and significant DMRs in 450k methylation array bulk breast cancer tissue relative to peripheral blood leukocytes (PBLs) (p < 0.0001 & absolute difference > 0.1). Top bar plots are colored by CpG islands (blue), shores (yellow), shelves (red) or open sea (brown) regions. Points in the plot below shows the observed z-score normalised overlapping count compared to the boxplots showing the distribution of overlapping counts between bulk breast tissue vs blood DMRs and random cfDNA background regions across 3000 permutations. Red points indicate significant overlap (p ≤ 0.01) in hypermethylated regions, while gray points indicate non-significant overlap (p > 0.01). **(C-D)** Overlap between the top 150 significantly hypermethylated pre-diagnosis cfDNA regions and significantly hypermethylated regions in **(C)** bulk breast cancer tissue versus blood, and **(D)** bulk breast cancer versus adjacent breast normal tissue. Boxplots represent distribution of overlap between significantly hypermethylated bulk breast cancer tissue markers and background cfDNA regions. **(E)** Intersecting regions significantly hypermethylated in bulk cancer, adjacent normal and healthy normal breast tissue relative to PBLs that overlap with 75 out of the top 150 cfDNA hypermethylated regions. (**F-G)** Overlap between 75 out of the top 150 hypermethylated pre-diagnosis cfDNA regions located in CpG islands and significantly hypermethylated regions

To further investigate the overlap in hypermethylated regions between cfDNA and hypermethylated regions in bulk breast tissue, we identified significant DMRs (FDR < 0.0001 & absolute methylation difference > 0.1) in 450k methylation array breast cancer tissue relative to PBLs. Among the 75 hypermethylated cfDNA regions, 47 (62.7%) regions overlapped with significantly hypermethylated regions in 450k methylation array bulk breast cancer tissue relative PBLs (Fig. 4B). To further evaluate whether the enrichment in hypermethylated regions between cfDNA and bulk breast cancer tissue methylation array profiles were significant, a permutation test was performed by comparing the observed number of overlapping regions to the expected overlap if random background cfDNA regions were selected. Most notably, only regions that were concordantly hypermethylated in pre-diagnosis cfDNA and in bulk breast cancer relative to PBLs tissue were significantly (p < 0.01) overlapping (Fig. 4B), whereas hypomethylated regions were not significantly enriched. Stratifying the enrichment by CpG island, shore, shelf, and open sea regions further revealed that the overlapping hypermethylated regions were most enriched among CpG islands, concordant with previous observations of hypermethylated regions in cancers being primarily observed in CpG islands (Fig. 4C) [36, 38]. Likewise, computing the overlap between the 75 hypermethylated cfDNA and significantly hypermethylated regions in bulk BRCA relative to ABRNM (Fig. 4D, Fig. S7E) and HBRNM (Fig. S7A & Fig. S8A) revealed 29 and 36 significantly overlapping regions respectively indicating that predictive cfDNA methylation markers consisted of regions uniquely hypermethylated in breast cancer tissue (Fig. 4E). To further ensure that the overlap wasn’t by chance or due to confoundment, as the bulk breast cancer and breast normal tissues methylation data were from separate studies, we recalculated the overlapping hypermethylated regions using bulk BRCA, ABRNM and HBRNM collected and processed from the same study (GSE69914). Consistent with the combined methylation array datasets, the cfDNA hypermethylated regions were also significantly overlapping with regions hypermethylated in BRCA relative to ABRNM (Fig. S7F & Fig. S8E) and HBRNM (Fig. S7G & Fig. S8F), particularly among CpG island regions.

We observed that many of the hypermethylated cfDNA regions mapped to promoter regions of tumour suppressor genes including GATA4, ZNF471 and SFRP, as well as in previously reported cfDNA breast cancer methylation markers (Fig. S9, Supplementary Table 4). Among proximal gene targets of hypermethylated cfDNA DMRs, various genes with dysregulated methylation were also inversely correlated with changes in expression between TCGA breast cancer and normal tissue among overlapping CpG sites, indicating that early changes in methylation detected from cfDNA may directly alter expression of these genes (Fig. S10). For example, ICAM2 expression has been implicated as a tumor suppressor that inhibits cancer cell invasion and migration, however we found an increase in methylation that directly correlated with a decrease in expression in breast cancer tissue [39]. Likewise, promoter methylation of genes such as *CDKL2* has been highlighted as a cfDNA methylation marker for triple negative breast cancers [15].

We further compared whether the identified overlapping hypermethylated cfDNA markers were specific to breast cancer tissue or potentially applicable to multiple cancers by calculating overlapping hypermethylated regions in pre-diagnosis cfDNA and in TCGA cancers across 13 different tissues relative to PBL methylation profiles (Fig. 4F & Fig. S11). Pre-diagnosis hypermethylated cfDNA markers in CpG island regions were most significantly enriched in breast tissue for both cancer and normal tissue relative to other tissue types (Fig. 4F-G), indicating that the hypermethylated CpG island regions detected in pre-diagnosis breast cancer cfDNA are likely specific to breast cancer tissue. However, we also observed significantly overlapping hypermethylated regions in other bulk cancer tissue types relative to PBL when including non-CpG island regions, which may suggest potential pan-cancer applications among captured markers.

## Discussion

Using cfMeDIP-Seq to profile cfDNA methylomes, we were able to capture cfDNA methylation signatures predictive of breast cancer development prior to clinical presentation and even in cases with a negative mammogram screen within a year before blood collection. We highlighted the predictive performance of using pre-diagnosis DMRs to classify individuals with underlying breast cancers within a discovery cohort and for predicting the absolute risk of an individual developing breast cancer within five years. We also demonstrate that these markers are detectable prior to mammogram detection and performs particularly well in detecting early breast cancers among women under the age of 50, for whom existing screening mammography programs are not universally recommended. Furthermore, we were able to validate the performance of top ranking hypermethylated regions by accurately discriminating a held-out batch of pre-diagnosis cases from controls at 53.3% sensitivity while retaining 100% specificity. Despite this, we acknowledge that the pre-diagnosis validation set sample sizes were small and additional validation of identified biomarkers will need to be further investigated in independent studies with larger cohorts. Our pre-diagnosis signatures are also highly generalizable to established metastatic breast cancers, providing further evidence that these signatures are related to the underlying malignancy. Likewise, the cfDNA methylation classifiers can also discriminate breast cancers from head and neck cancers cases, although follow-up investigations will be necessary to assess the specificity of our markers as they relate to cancer type and histology. We found that among the top 150 cfDNA hypermethylated regions detected in the discovery set, hypermethylated cfDNA regions at CpG island were the most significantly enriched for in breast cancer tissue relative to PBLs and normal breast tissues, revealing that the detected cfDNA hypermethylated regions captured in pre-diagnosis samples are partially reflective of breast cancer methylomes. However, it should be noted that among the top 150 hypermethylated regions identified from pre-diagnosis cases, only 62.7% of regions were concordantly hypermethylated in breast cancer tissue, which may implicate other non-breast cancer tissue markers being predictive of breast cancer risk among remaining non-overlapping regions. It is likely that other phenotypes such as paraneoplastic syndrome, changes in the tumour microenvironment or other changes in immune profiles, may also contribute, and improve risk prediction. Additional investigations deconvoluting immune profiles and other tissue types from the detected cfDNA methylation markers will be needed to further investigate the sources of non-overlapping regions.

There has been an increasing consensus among recent studies that cfDNA methylation profiles, often combined with other biomarker or imaging-based approaches, can yield the best predictive performance for detecting cancers at early stages [12]. However, to implement liquid biopsies for population screening of cancers, the viability of existing assays and predictive models needs to be demonstrated in biologic specimens collected prior to a cancer diagnosis. Our work builds on major investments made to establish large longitudinal population cohorts that store samples collected from healthy individuals at the time of study recruitment. By linking participants to administrative data routinely collected in public health settings in Canada, we can follow up and identify the occurrence of morbidities such as cancers. These types of cohort resources allow for interrogation of pre-diagnosis biologic samples, as we described here using developments in cfDNA methylation profiling assays and can be similarly extended to other cancers, as demonstrated using OHS incident prostate cancer samples [40], and alternative emerging methodologies interrogating blood biomarkers such as cell-free RNA, proteins, and metabolites.

Several recent studies have profiled plasma cfDNA methylation profiles of breast cancers for early cancer detection, however these studies are primarily sampled from patients after clinical detection or formal diagnoses, and typically use a pre-designed enrichment panel to target specific genomic loci [13-16]. To date, only one study has profiled breast cancer plasma collected prior to clinical detection: in that investigation using a single methylome marker, reported sensitivities were between 5-12% with 88% specificity among samples collected two to three years before diagnosis [41]. Comparatively, our predictive models achieve a mean sensitivity of 60% at 100% specificity for classifying test set breast cancer cases diagnosed up to six years following blood plasma profiling. Acknowledging the sample size of pre-diagnosis test samples was small, additional independent studies are needed to further validate the utility of cfDNA methylation markers in pre-diagnosis samples. While we reported higher sensitivity for HR positive breast cancer in both the discovery and late-stage breast cancer samples, existing methylome profiling of plasma samples collected at the time of cancer diagnosis, and presumably more advanced breast cancer patients have typically noted better classification performance among HR negative breast cancers relative to HR positive [15, 42]. As the majority of breast cancers are HR positive, the incidence rate of HR negative breast cancer in the OHS is relatively low, and we suspect a poorer predictive performance among HR negative breast cancers may arise from biasing toward selected features associated with the more numerous HR positive breast cancers. Alternatively, considering that HR positive breast cancers typically have slower doubling times, less aggressive cancers may be present for longer but remain undetected by mammograms until reaching visible sizes allowing for a longer window of opportunity for detection at an early stage and age. Conversely, aggressive cancers which develop and expand more rapidly, may have a shorter window of opportunity for detection at an early stage.

The batching of case and control groups during sequencing are often not reported across early cancer detection studies. Unfortunately, internal model performance can often be inflated if cases and controls are processed in separate batches. When case and control groups are perfectly confounded between batches, signals associated with technical artifacts can often drive separation of case and control groups in both training and testing samples, consequently conflating predictive performances [29, 43]. Accordingly, we profiled our cases with control samples between sequencing runs in this study, in addition to using a repeated cross-validation approach to estimate the uncertainty of predictive performances. Likewise using held-out a batch of pre-diagnosis cases and control samples, we demonstrate that the captured markers are predictive of breast cancer in an independent set of samples, and similarly demonstrate that the developed classifiers were also highly robust for the identification of established breast cancers from a separate cohort. False-positive predictions in our cohort may still represent misclassifications of control samples with undetected underlying cancers, owing to variable follow-up duration among cancer-free controls (Supplementary Table 2).

Additionally, the following limitations of the current study should be considered when interpreting our findings. Firstly, 1.6 mL of plasma was used per participant for this study, which is a substantial amount of biobanked material, but larger plasma volumes would likely increase the number of ctDNA fragments captured and further improve detection sensitivity. Owing to the prospective nature of the OHS cohort, our sample sizes of pre-diagnosis cancers were limited by the incidence rate of the cancer among the study population with a cryopreserved blood sample, acknowledging that these incident cases will accrue with time. Additionally, not all cancer-free control samples were followed up for the same duration owing to our matching of controls to cases by sample collection time. While we followed all controls up to 2019 to ensure that they were alive and free of cancer, it is possible that controls with shorter follow up times may have underlying undetected cancers that had yet to be diagnosed as suggested by the lack of high risk samples assigned in the high risk group according to classification scores in either the discovery or validation sets. Consequently, this may inflate the false positive rate by mislabelling control samples with undiagnosed cancers. Likewise, the false negative rate may also be inflated by reducing the power for detecting cancer specific DMRs if controls with undiagnosed cancers harbored the same hypermethylated regions with pre-diagnosis cases.

## Conclusions

Despite the current limitations described above, genome-wide cfDNA methylation profiling of pre-diagnosis breast cancer plasma samples reveals detectable signatures that are predictive of breast cancer risk up to six years prior to diagnosis. Currently, breast cancer is one of the few cancer types with an established population screening tool owing to its associated reduction in mortality. Consequently, most breast cancers are typically diagnosed at stage I or II as seen in the OHS cohort and across the population. While mammograms are currently the gold standard for early breast cancer screening achieving a 92% sensitivity and 92% specificity in Ontario [44], low adherence to screening guidelines is recognized, substantial physical and personnel resources are required to deliver such screening, and low-dose radiation exposure may also increase the risk of future breast cancer development [45]. A liquid biopsy-based approach could not only enable simultaneous detection of multiple cancer types, but also mitigate risks associated with radiographic imaging approaches, and be relevant to groups of individuals where mammographic screening methods are not currently recommended. While it is unclear whether diagnoses prior to mammographic detection will further improve prognostic outcomes, detection of breast cancer signatures up to six years prior to a stage I or II diagnosis presents potential opportunities for early pre-symptomatic detection and intervention among other cancer types with no reliable screening tool. While sample sizes of our validation sets are limited in our study, we find that methylation signatures concordant with breast cancer tissue can be detected in cfDNA preceding mammogram detection. Indeed, future applications of liquid biopsies for early cancer detection will require identifying the tissue of origin of underlying cancers. Likewise, profiling of pre-diagnosis plasma from individuals with other cancer types will allow for identifying tissue-specific markers and the development of tissue of origin classifiers, similar to those demonstrated in existing studies classifying samples with established cancers.

## Data Availability

The datasets used and/or analysed during the current study are available from the corresponding author on reasonable request.

## Abbreviations

ABRNM: Adjacent Breast Normal
AUC: Area Under the Receiver Operating Characteristic Curve
AUC(t): Time-dependent Area Under the Receiver Operating Characteristic Curve
HBRNM: Healthy Breast Normal
BRCA: Breast Cancer
cfDNA: Cell-free DNA
C-index: Concordance Index
CoxPH: Cox Proportional Hazard
CV: Cross-validation
DMR: Differentially Methylated Regions
GEO: Geo Expression Omnibus
IC: Input Control Library
IHEC: International Human Epigenetics Consortium
IP: Input Library
KM: Kaplan-Meier
LogFC: Log Fold-Change
OCTANE: Ontario-wide Cancer TArgeted Nucleic Acid Evaluation
OHS: Ontario Healthy Study
TCGA: The Cancer Genome Atlas

## Declarations

### Ethics approval and consent to participate

Patient plasma samples were obtained from the Ontario Health Study (OHS) with protocols approved by the University of Toronto Health Sciences research ethics board (protocol #34088). All participants gave written informed consent prior to participation. All samples and participant data were deidentified and assigned unique research IDs. Original OHS participant and CCO IDs are not known to anyone outside the research group. Supplementary tables do not contain any information that enables identification of the original participants.

## Acknowledgments

We would like to thank the Genomics Research Platform team at OICR for performing the cfMeDIP-Seq assay on the plasma samples, as well as the insightful comments on the study from members of the Ontario Institute for Cancer Research and Princess Margaret Cancer Centre. We would also like to thank OCTANE investigators and study staff. Parts of the material are based on data and information provided by Ontario Health, and includes data received by Ontario Health from the Canadian Institute for Health Information (CIHI) and the Ministry of Health (MOH). Funding was provided an Adaptive Oncology grant from the Ontario Ministry of Universities and Colleges. The opinions, reviews, views and conclusions reported in this publication are those of the authors and do not necessarily reflect those of Ontario Health, CIHI, and/or the MOH. No endorsement by Ontario Health, CIHI, and/or the MOH is intended or should be inferred.

## Author Contributions

PA conceived the project. PA and KS are responsible for funding acquisition. KS performed administrative linkage to identify incident cancer cases in the OHS. PA, NC and DS conceptualized the project and study design. NC, PA and DS designed the computational and statistical analyses. NC performed the bioinformatics, statistical analysis, figure generation and manuscript writing. AS performed the bioinformatics for external validation samples. DWC and SVB provided external validation data. PA, DS, SVB and DDC advised on the computational and statistical analyses. PA, DS, SVB, DWC, KS, TWO wrote and revised the manuscript.

---

**Fig. S1:**
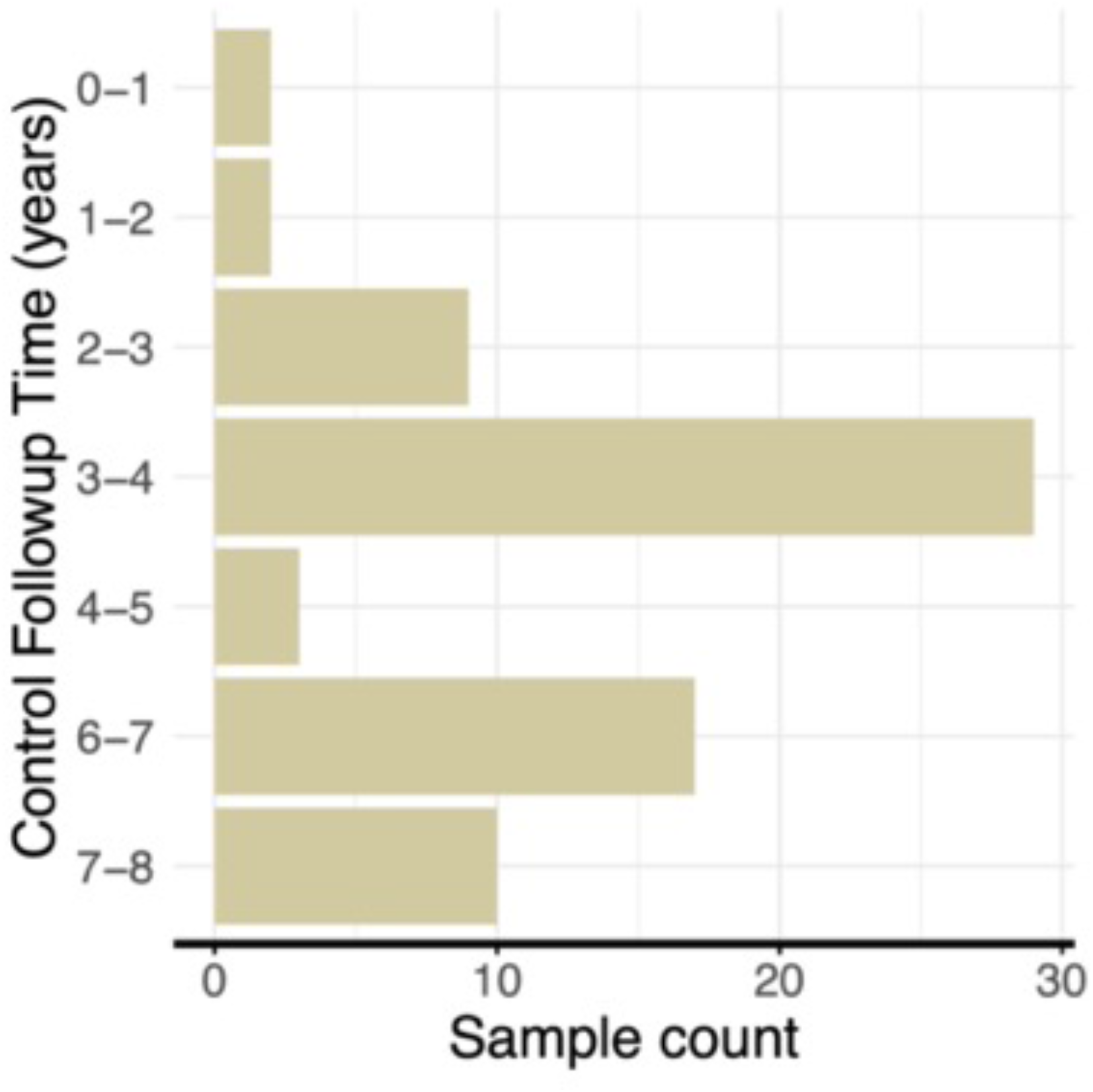
Matched cancer-free control sample follow-up time. Selected control plasma (n = 70) were matched to cases by sex, age, time of sample collection, smoking status and alcohol consumption frequency.

**Fig. S2:**
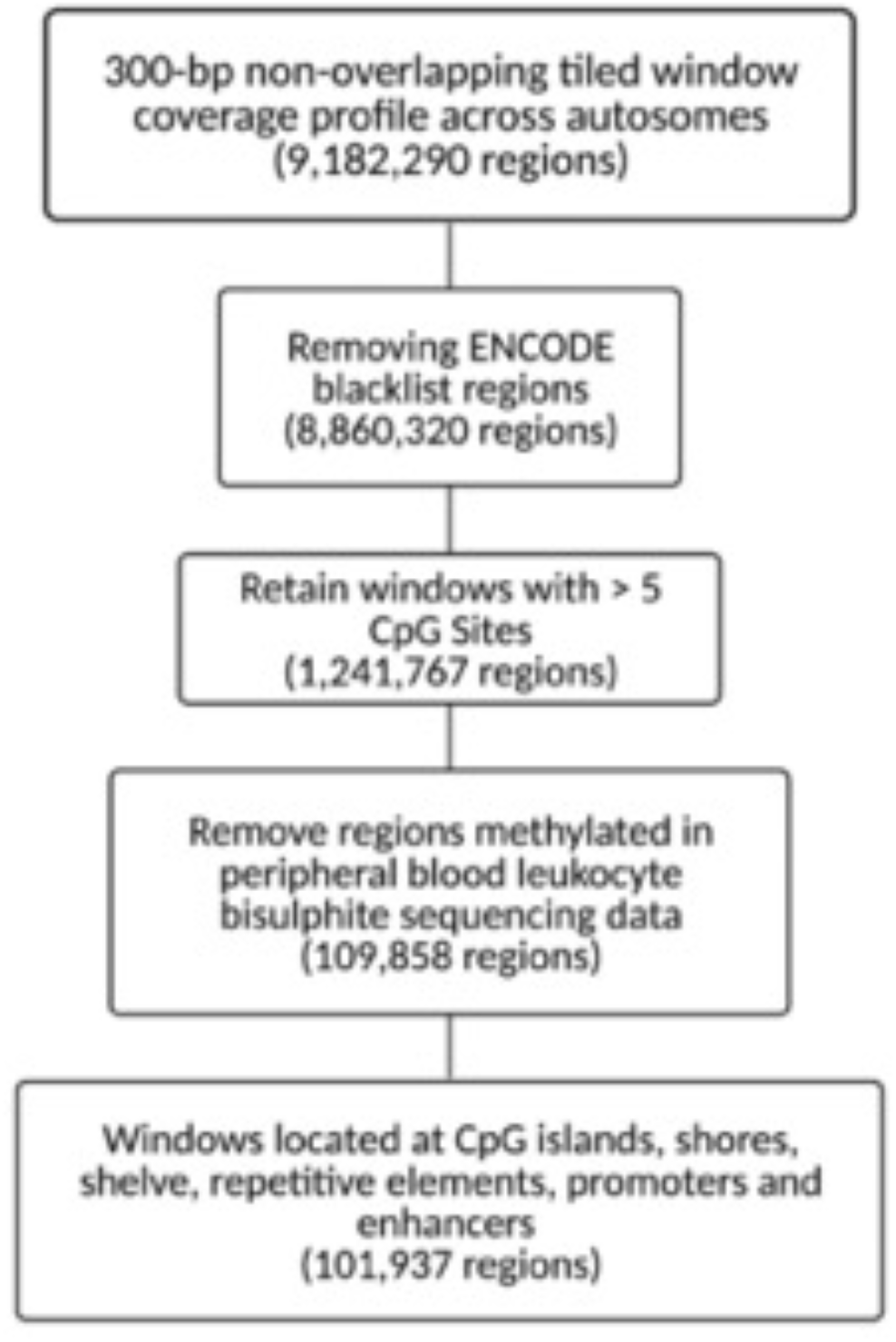
Flowchart of filters applied to reduce background regions methylated and enrich for DMRs associated with breast cancer. Flowchart of filters applied to reduce background regions methylated in peripheral blood leukocytes and enrich for DMRs associated with breast cancer development. Feature search space started with genome-wide coverage profiles across 9,182,290 300-bp nonoverlapping tiled windows. To reduce background signals derived from peripheral blood cells (PBLs) and enrich for tumour-derived signals, regions frequently methylated in PBLs (average methylation across 300 base pair window of over 0.25) whole-genome bisulfite sequencing data from IHEC were filtered out (n = 79). To enrich for CpG dense and regulatory regions, windows with at least six or more CpG sites, and located at CpG islands, shores, shelves, repeat elements, and FANTOM5 enhancers or promoters were selected, leaving 101,937 regions remaining to perform differential methylation analysis.

**Fig. S3:**
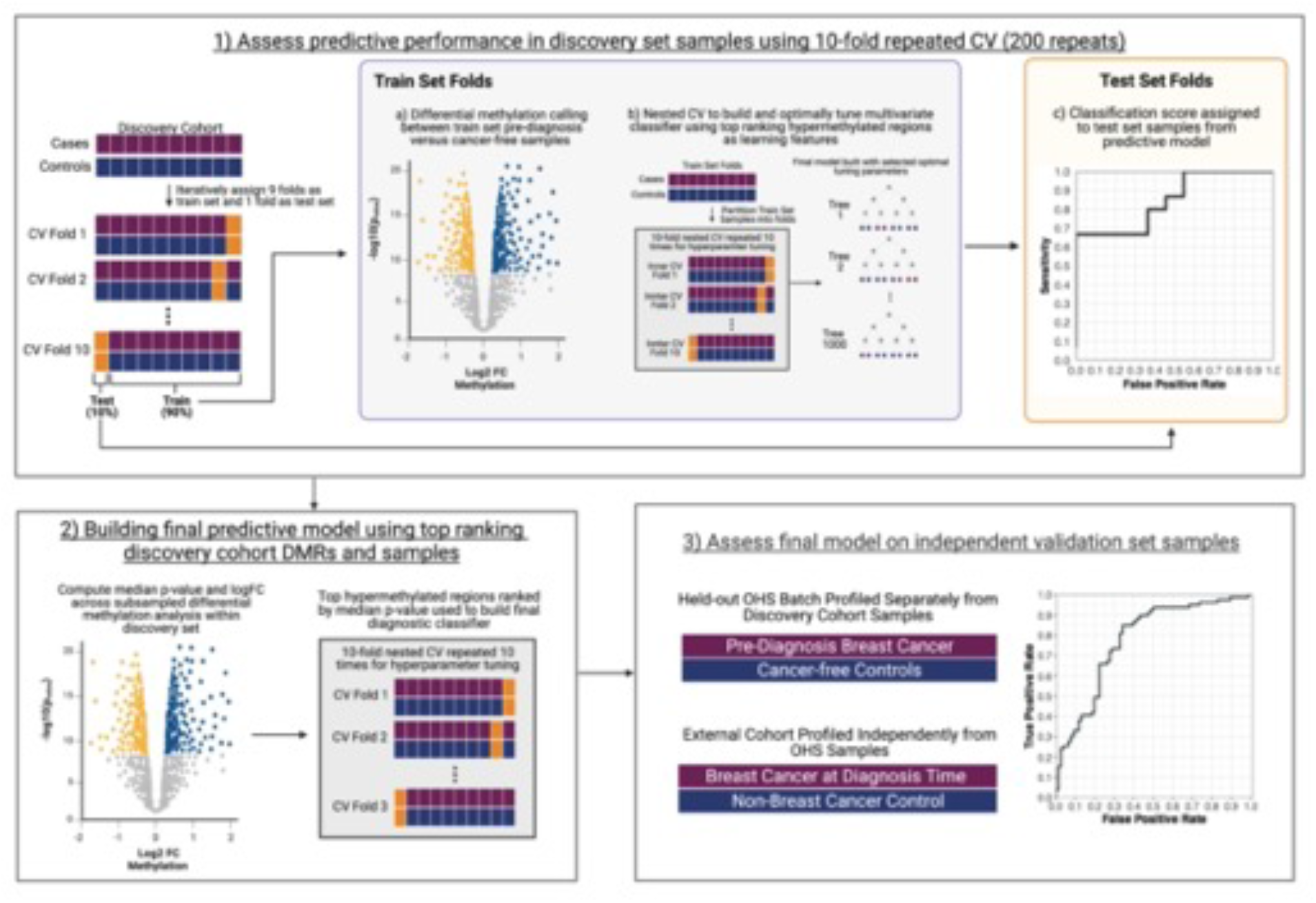
Schematic of the analytical approach performed to assess predictive performance of pre-diagnosis cfDNA methylation profiles. To assess predictive performance in discovery cohort samples and identify optimal number of features to use in the final classifier, a 10-fold CV repeated 100 times was performed on the discovery set. Pre-diagnosis breast cancer cases and controls were partitioned into 10-fold splits, splitting the number of cases by years prior to diagnosis evenly among each fold. Iteratively, nine-folds were selected as train set samples and used to perform differential methylation calling to identify and rank the top hypermethylated regions in pre-diagnosis breast cancer cfDNA. The top ranking hypermethylated regions among pre-diagnosis cases in the 9-folds are used to build a random forest diagnostic classifier to discriminate individuals with undetected breast cancers. Samples in the one remaining held-out test fold that was not involved in any aspect of feature selection or model building were assigned a classification score from the model built using nine train folds to evaluate the classifier performance. This process was iteratively repeated 10 times, such that each fold was the held-out test fold once. We repeated this 10-fold CV split strategy 100 times, each time with random sample partitioning into folds to infer the overall performance in discovery cohort samples. Across the 100 repeats, differential methylation calling was performed 1000 times (10 times per CV repeat) with different subsampled cases and controls. The mean logFC in methylation and p-value was computed for each region, and ranked according to p-value. The top 150 hypermethylated regions were used to build diagnostic classifiers trained with all discovery cohort samples, using a 10-fold nested CV repeated 10 times for hyperparameter tuning. The classifier performance was evaluated on independent test set samples consisting of a held-out batch of pre-diagnosis cases and controls from OHS and stage IV breast cancers collected at the time of diagnosis and controls from external cohorts.

**Fig. S4:**
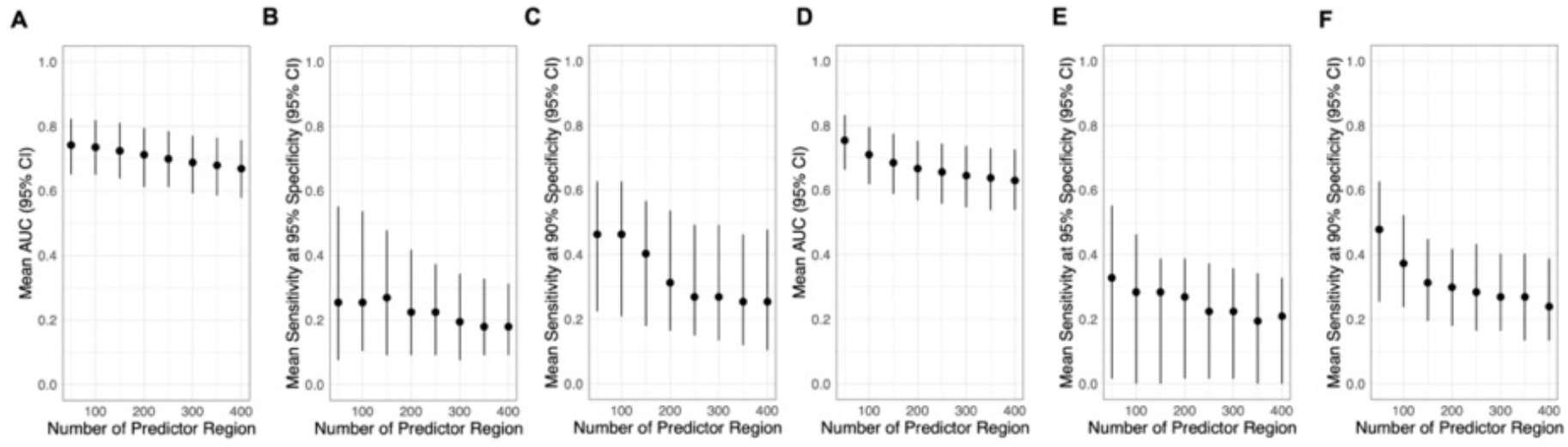
Discovery cohort test-fold performance across varying number of top ranking regions. Across the 100 repeated 10-fold cross validation, the top hypermethylated features identified from train set folds were ranked according to p-values calculated by performing **(A-C)** a Wald’s test of the negative binomial regression coefficient between pre-diagnosis cases and controls and **(D-F)** a Bartlett’s test for variability between pre-diagnosis cases and controls within each 10-fold CV repeat. Diagnostic classifiers built using the top ranking features in train set folds were assessed on held-out test folds by assigning classification scores. Average test-fold classification scores were computed for each sample across the 100 repeats. Averaged risk scores were bootstrapped 1000 times to obtain **(A & D)** mean AUROC, **(B & E)** mean sensitivity at 95% specificity and **(C & F)** sensitivity at 90% specificity. Dots indicate mean performance and lines represent 95% percent confidence intervals.

**Fig. S5:**
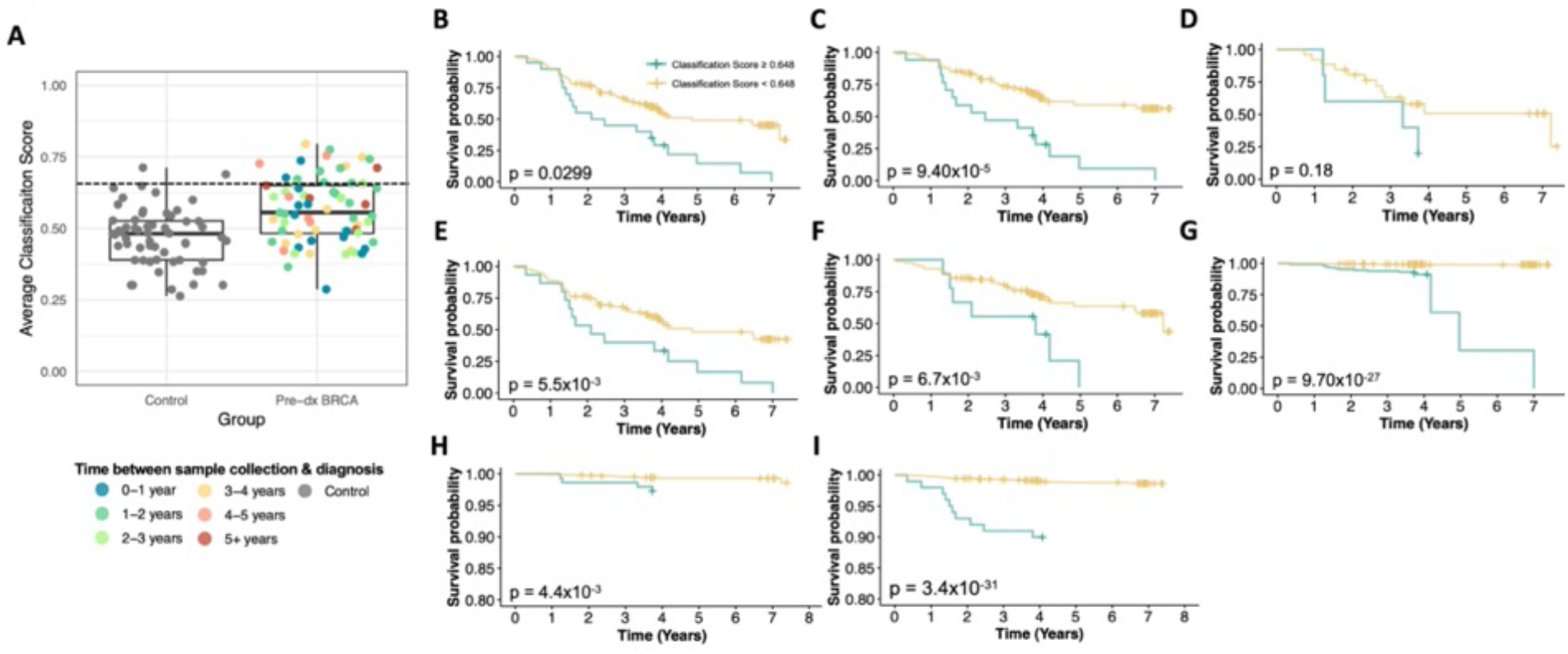
Kaplan-Meier survival curves of discovery set samples stratified by mean test-fold classification scores. **(A)** Average classification scores for controls and pre-diagnosis breast cancer (BRCA) cases in the discovery cohort. Mean classification scores were calculated by averaging all test-fold classification score across repeats per sample. Each dot represents a sample colored by the time between sample collection and diagnosis for cases and grey for controls. Dotted line indicates classification score (0.648) yielding 95% specificity. **(B-F)** Unweighted Kaplan-Meier survival curves stratified by discovery cohort samples above and below a mean classification score of 0.648 for controls and **(B)** all cases, **(C)** hormone receptor positive cases, **(D)** cases diagnosed between ages 31-50, **(E)** cases diagnosed between ages 51-75, and **(F)** cases with a negative mammogram screen within one year of diagnosis. **(G-I)** Kaplan-Meier survival curves weighted by age specific cumulative breast cancer incidence rates from the Canadian Cancer Registry stratifying controls and **(G)** hormone receptor positive cases, **(H)** cases diagnosed at ages 31-50, and **(I)** cases diagnosed at ages 51-75.

**Fig. S6:**
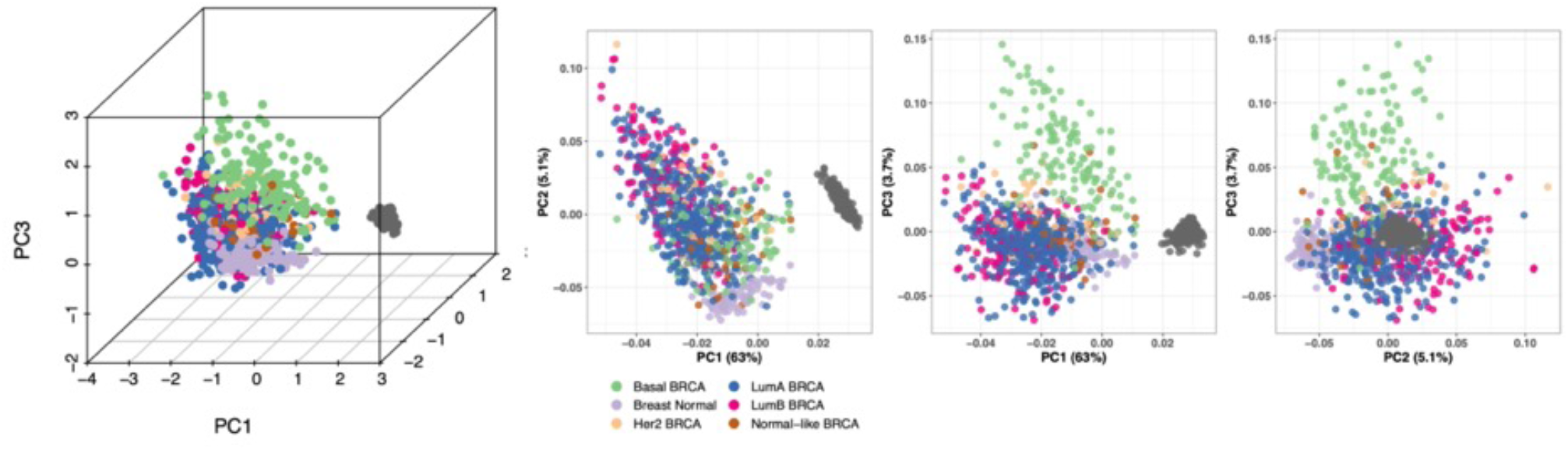
Pre-diagnosis cfDNA methylation signatures discriminates bulk breast cancer. Principal component analysis of TCGA 450k methylation array of bulk breast cancer tissue (n = 787), bulk normal breast tissue (n = 97) and peripheral blood leukocytes (n = 628) across 156 CpG sites overlapping the 75 out of the top 150 hypermethylated regions in pre-diagnosis breast cancer cfDNA that contain at least one CpG site profiled by the 450k methylation array.

**Fig. S7:**
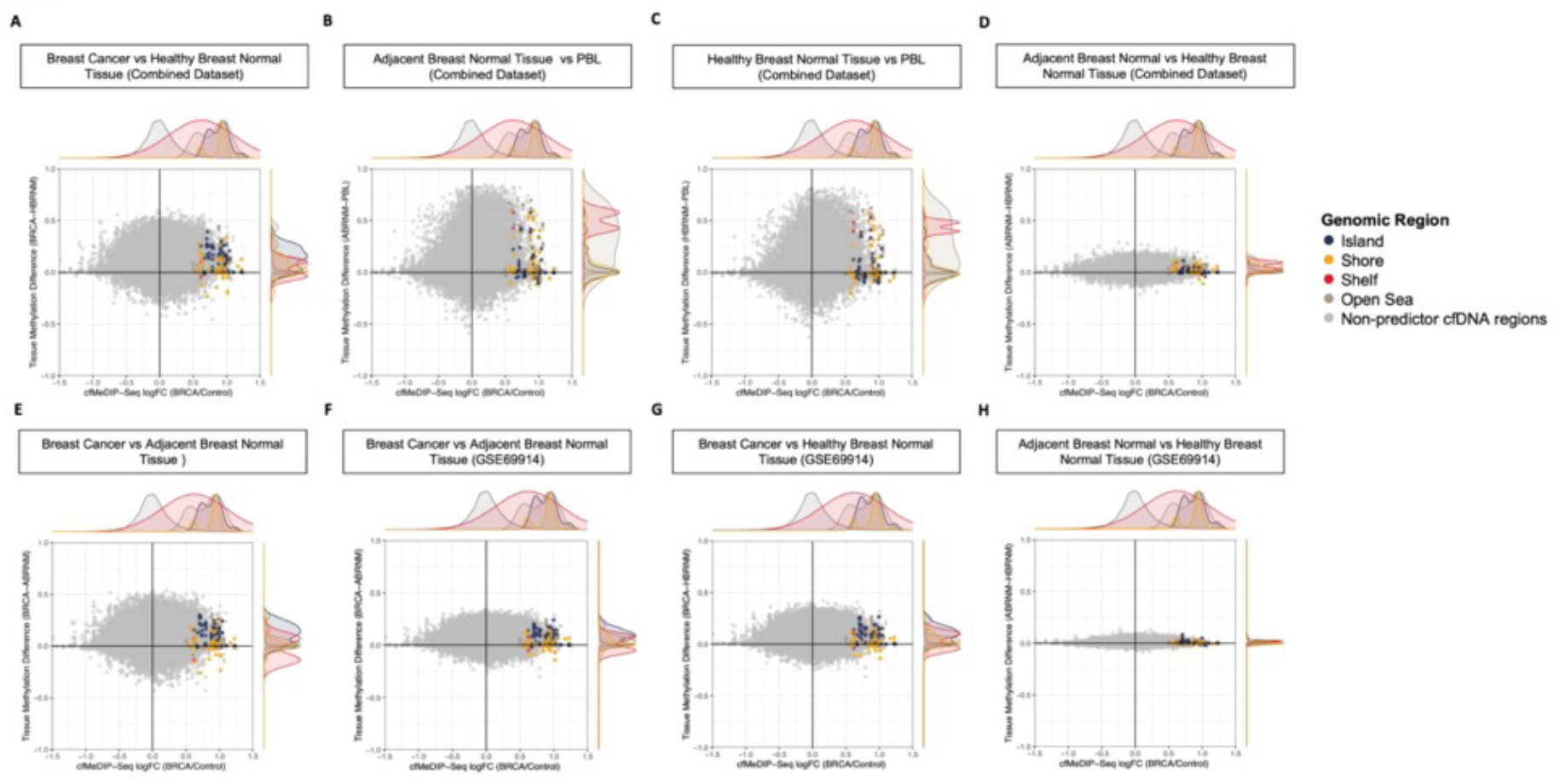
Association between cfDNA methylation and in bulk breast tissue methylation profiles. **(A-E)** Log-fold change in cfDNA methylation between cases and controls in background (grey) and in 75 regions out of the top 150 hypermethylated (colored) regions identified from discovery cohort pre-diagnosis breast cancers targeted by the 450k DNA methylation array compared to the absolute change in methylation in overlapping sites between bulk **(A)** breast cancer (BRCA) tissue vs healthy breast tissue (HBRNM), **(B)** adjacent breast normal tissue (ABRNM) vs peripheral blood leukocytes (PBL), **(C)** HBRNM vs PBLs, **(D)** ABRNM vs HBRNM, and **(E)** BRCA vs ABRNM in combined 450k methylation array datasets (GSE133985, GSE74214, GSE88883, GSE66313 & GSE101961). **(F-H)** Log-fold change in methylation in cfDNA compared to the absolute change in methylation among overlapping CpG sites between **(F)** BRCA vs ABRNM, **(G)** BRCA vs HBNM and **(H)** ABRNM vs HBRNM in 450k methylation array data from GSE69914. Each point represents a CpG site on the 450k methylation array and colors indicate the genomic region the CpG site is located for predictor regions.

**Fig. S8:**
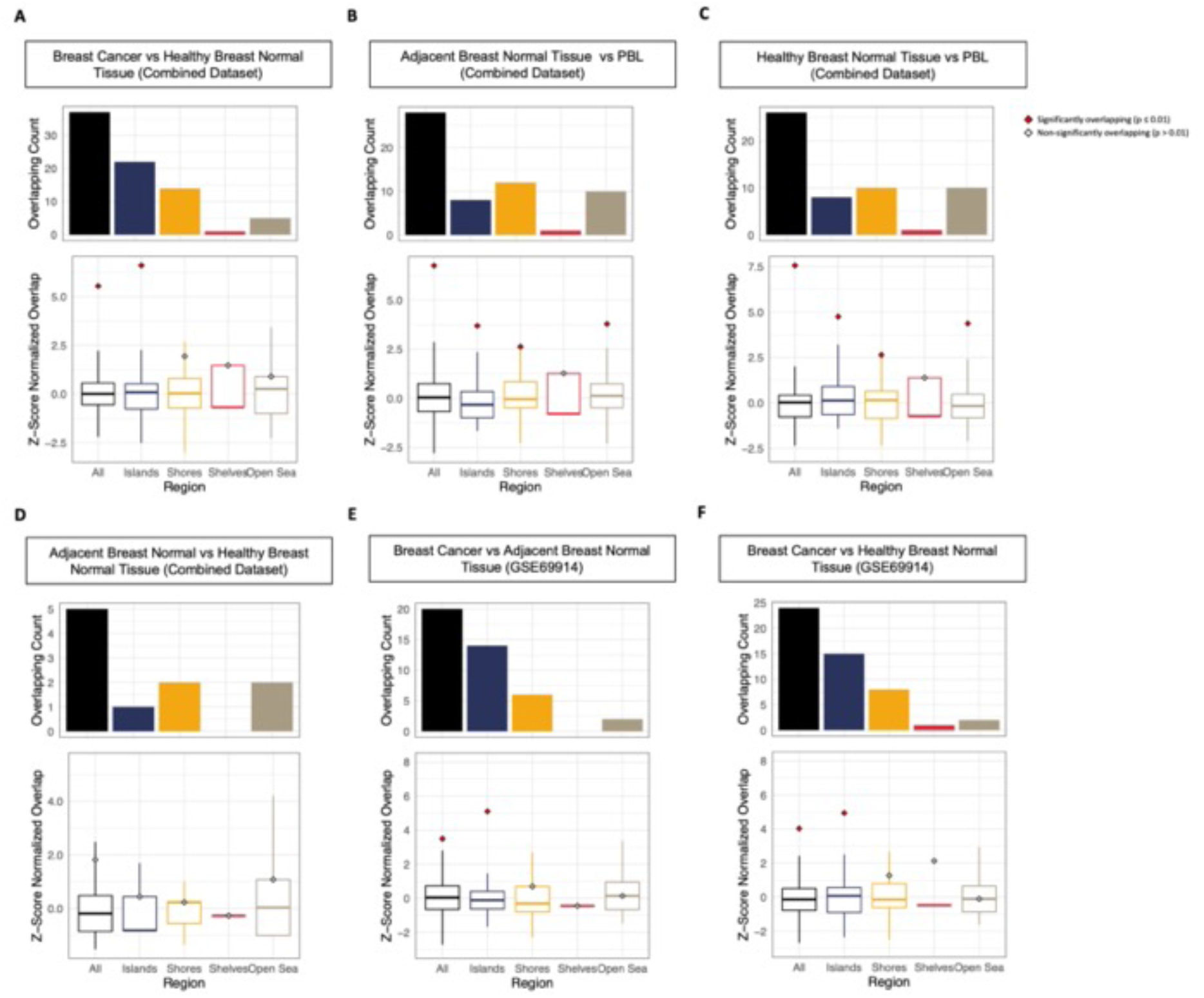
Overlapping hypermethylated regions between pre-diagnosis cfDNA and bulk breast tissue samples. **(A-D)** Observed overlapping hypermethylated regions among predictor cfDNA regions and significantly hypermethylated CpG sites (absolute difference > 0.1 & p < 0.0001) between **(A)** BRCA vs HBRNM, **(B)** ABRNM vs PBLs, **(C)** HBRNM vs PBLs **(D)** ABRNM vs HBRNM in combined methylation array datasets (GSE133985, GSE74214, GSE88883, GSE66313 & GSE101961). **(E-F)** Observed overlap between **(E)** BRCA vs ABRNM and **(F)** BRCA vs HBRNM in 450k methylation array data from GSE69914. Bar plots represent observed number of overlapping regions between cfDNA and bulk tissue, while significance of the overlap was determined through a permutation test comparing the overlap between background regions in cfDNA and significantly hypermethylated regions bulk tissue profiles (repeated 3000 times with random sets of subsampled cfDNA regions). Counts from background overlap are z-score normalized and shown in the boxplots. Points indicate the observed z-score normalized overlap with red indicating a significant overlap (p<0.01) while gray points indicates non-significant overlaps (p > 0.01).

**Fig. S9:**
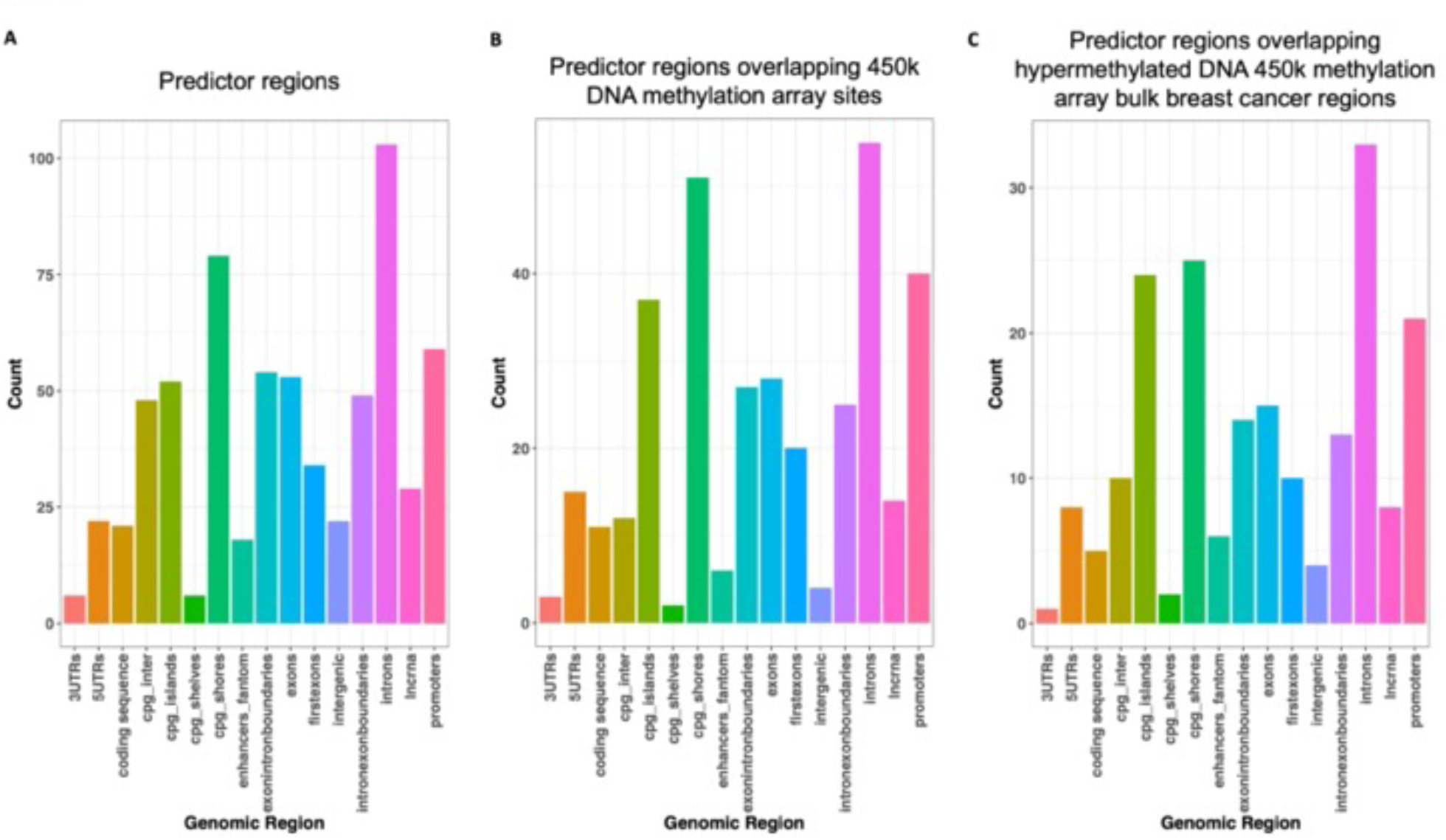
Genomic location of predictive pre-diagnosis cfDNA hypermethylated regions. Proportion of hypermethylated regions across genomic annotations in **(A)** The predictor regions comprised of the top 150 hypermethylated windows in discovery cohort, **(B)** predictor regions overlapping sites profiled by 450k DNA methylation array and **(C)** predictor regions overlapping sites profiled by 450k DNA methylation array that is also hypermethylated in bulk breast cancer tissue relative to PBLs or breast normal tissue.

**Fig. S10:**
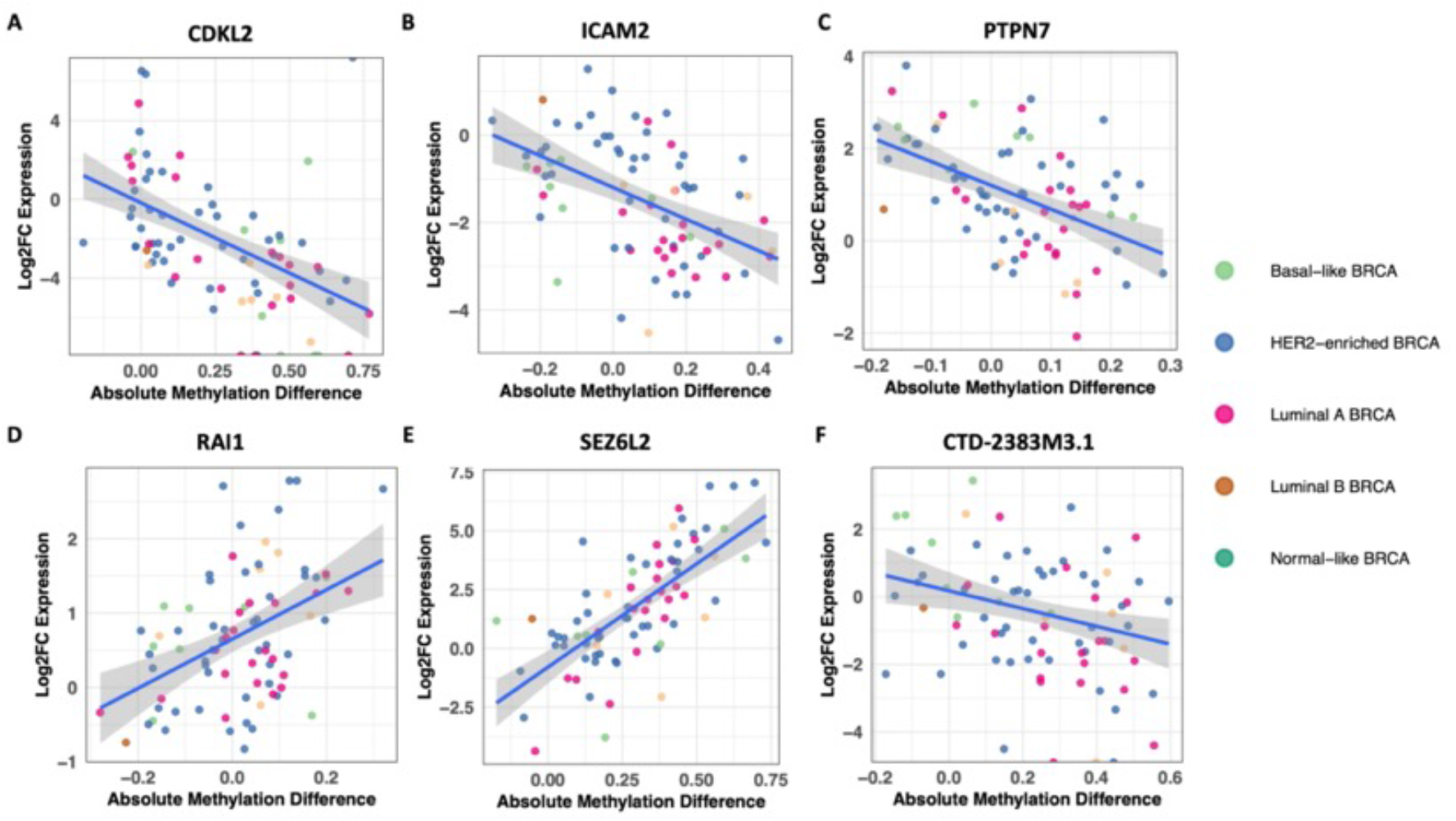
Hypermethylated regions in cfDNA associated with a change in gene expression. Absolute change in promoter methylation and log2 fold-change in gene expression between TCGA breast cancer and adjacent breast normal tissue among the top 150 hypermethylated regions pre-diagnosis breast cancer cfDNA for **(A)** CDKL2, **(B)** ICAM2, **(C)** PTPN7, **(D)** RAI1, **(E)** SEZ6L2, **(F)** CTD-2383M3.1.

**. S11:**
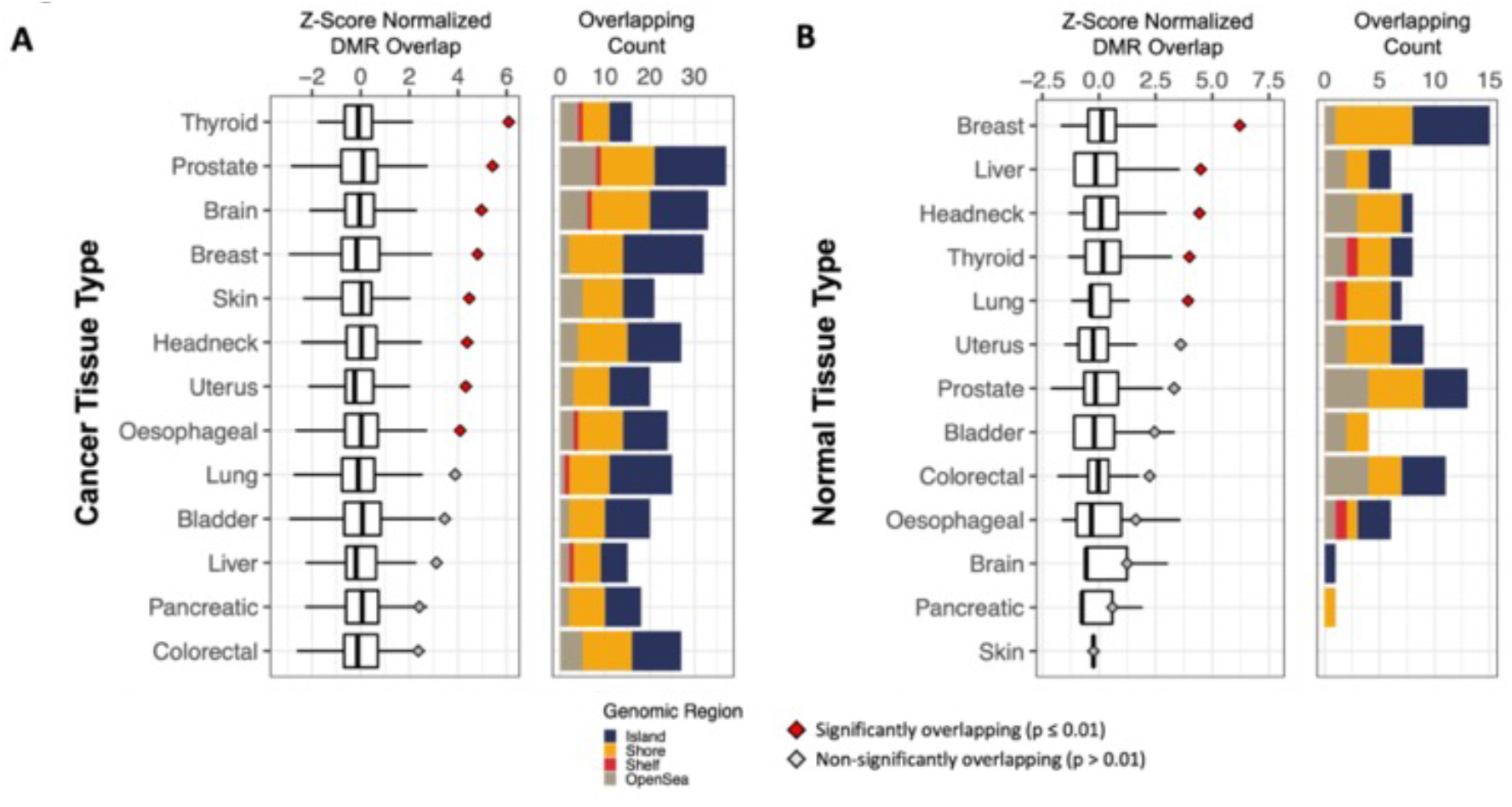
Overlapping hypermethylated regions between cfDNA and bulk cancer and normal tissue relative to PBLs. Differentially methylated CpG sites were identified between TCGA bulk adjacent normal versus PBLs for each tissue type using a standard F-test. Bar plots show the overlapping count of significantly hypermethylated regions (absolute difference > 0.1 and q-value < 0.001) in **(A)** bulk cancer tissue vs PBLs, and **(B)** bulk normal tissue vs PBLs that overlap with the top 150 cfDNA hypermethylated regions. Barplots are colored by genomic regions with blue representing CpG islands, yellow as shores, red as shelves and brown as open sea regions. The observed overlap was compared to the expected number of overlaps by computing the overlap between significant bulk tissue hypermethylated regions and randomly subsampled background cfDNA regions repeated 3000 times. The expected overlap are shown in box plots for 3000 random subsampling iterations following z-score normalization, while the points illustrate the observed z-score normalized overlap with cfDNA hypermethylated regions. Red points indicate significant (p < 0.01) overlap between cfDNA hypermethylated regions and bulk tissue hypermethylated regions, while gray points indicate non-significant overlaps.

**Supplementary Table 1: Baseline characteristics of pre-diagnosis cases and cancer-free control samples at time of blood collection**. Summary of age, sex, time since blood plasma collection, last mammogram prior to blood collection, ethnicity, body mass index (BMI), smoking frequency and alcohol consumption frequency among discovery and validation breast cancer cases and controls.

**Supplementary Table 2: Quality control, clinical and additional participant information across OHS samples**. Information across individual samples indicating age, CpG enrichment scores, methylated spike-in read proportions out of all methylation and non-methylation spike-in reads (thaliana beta), total reads following UMI deduplication, sample group, sample age, time between sample collection and diagnosis for cases, follow up time among controls, hormone receptor status of breast cancers and stage at diagnosis.

**Supplementary Table 3: Breast cancer cumulative incidence in OHS**. Cumulative incidence is presented as a function of time given a high (> 0.648) or low (< 0.648) classification score. Observed (Kaplan Meier) and predicted (Cox PH) probabilities are given stratified by risk score groups in discovery and validation samples.

**Supplementary Table 4: Genomic annotations across the top 150 hypermethylated predictor regions in pre-diagnosis cfDNA discovery set samples**. Genomic annotations performed using the Annotatr R package. Regions are also annotated to inform whether the region overlaps with sites on the 450k DNA methylation array and in hypermethylated bulk breast cancer tissue relative to PBLs and breast normal tissues.

